# InterMob: A 24-month randomised controlled trial comparing the effectiveness of an intervention including behavioural change techniques and free transport versus an intervention including air pollution awareness-raising on car use reduction among regular car users living in Grenoble, France

**DOI:** 10.1101/2022.07.21.22277902

**Authors:** Claudia Teran-Escobar, Sarah Duché, Hélène Bouscasse, Sandrine Isoard-Gatheur, Patrick Juen, Lilas Lacoste, Sarah Lyon-Caen, Sandrine Mathy, Estelle Ployon, Anna Risch, Philippe Sarrazin, Rémy Slama, Kamila Tabaka, Carole Treibich, Sonia Chardonnel, Aïna Chalabaev

## Abstract

**Background:** Frequent car use contributes to health and environmental issues such as air pollution, climate change and obesity. Active and sustainable mobility (bike, walk, public transport, car sharing) may address these issues. Different strategies have been implemented in past research, involving *hard* levers, aimed at modifying the economical or geographical context (e.g., free public transport), and *soft* levers, aimed at modifying psychological processes (e.g., personalised transport advice). However, few studies have combined both hard and soft levers. In addition, few have used robust methodologies (e.g., randomised controlled trials), followed behavioural changes in the long-term, and been anchored in behaviour change theories. InterMob aims to address these limits by implementing a 24-month randomised controlled trial including *hard* and *soft* levers. The objectives of InterMob are to a) evaluate the effectiveness of an experimental arm versus an active controlled arm, and b) identify the processes of mobility change.

**Methods:** Regular car users living in Grenoble (*N* = 300) will be recruited and randomised to one of the two arms. The experimental arm consists in a six-month intervention combining *hard* levers (free access to transport/bikes), and *soft* levers (e.g., personalised transport advice). The control arm consists in a six-month intervention aimed at raising awareness on air pollution and its health effects. Both arms will include eight evaluation weeks (spread out over 24 months) based on a GPS, an accelerometer, and a pollution sensor. Moreover, participants will complete mobility logbooks and surveys measuring psychological constructs, socio-economical, and socio-spatial characteristics.

**Discussion:** InterMob will assess the effectiveness of two interventions aimed at reducing car use within regular car users in the short-, mid- and long-term. Moreover, InterMob will allow to better understand the psychological processes of behaviour change, and the socio-economical and geographical conditions under which the intervention is efficient in reducing car use. Finally, the benefits of mobility change in terms of physical activity, quality of life, and exposure to pollution will be quantified.

**Trial registration:** ClinicalTrials.gov: NCT05096000

## Background

Daily car use contributes to major health and environmental issues. Motorised transport represents an important source of air pollution, noise and greenhouse gases (CITEPA, 2020; van Schalkwyk & Mindell, 2018; WHO, 2018). Moreover, regular car users present lower levels of physical activity (Chakrabarti & Shin, 2017), spend more time in sedentary behaviours (Sugiyama et al., 2012), and have a higher risk of being obese or overweight (Frank et al., 2004; Jacobson et al., 2011). In turn, air pollution, climate change and physical inactivity represent important dangers to human health and life expectancy (Fuller et al., 2022; Guthold et al., 2018; Kohl et al., 2012; Manisalidis et al., 2020). Nevertheless, despite all the negative consequences of daily car use, the car remains the main mode of transport in most countries (e.g., in France, 63% of daily trips are made by car, SDES, 2020).

Active and sustainable mobility such as cycling, walking, public transport and carpooling have in contrast positive effects on health and well-being. These modes of transport are associated with higher levels of physical activity (Chaix et al., 2014, 2019; Pucher et al., 2010; Rojas-Rueda et al., 2011), greater well-being (Martin et al., 2014), better work performances (Ma & Ye, 2019), and a higher life expectancy (Cepeda et al., 2017). In addition, these transport modes might represent an effective lever to tackle air pollution and climate change (Bernard et al., 2021; Brand et al., 2021). Nevertheless, active and sustainable mobility are less frequently used than the car for daily mobility (i.e., in France, 37% of daily trips are made by using an active and sustainable mobility, SDES, 2020).

### 1.1 Strategies for car use reduction, active and sustainable mobility promotion

During the last years, many interventions have been developed to reduce car use and promote active and sustainable mobility. They can be distinguished based on whether they involve *hard* or *soft* levers (Bamberg et al., 2011).

*Hard* levers are strategies that target a change in the geographical and economic context of individuals to encourage them to reduce car use and/or increase active mobility. Hard-lever interventions include for example the implementation of low-emission zones (i.e., restricting the entry of polluting motorised transport into an established area), free public transport, city tolls or new cycling and walking infrastructures (Gärling & Schuitema, 2007; Kuss & Nicholas, 2022; Martin et al., 2012; Mölenberg et al., 2019). Results indicate that in general, *hard* levers seem to increase the use of active and sustainable mobility and reduce car use (e.g., implementing a payment for entering to the city centre reduces between 12 and 33% of city-centre cars in European cities, Kuss & Nicholas, 2022; infrastructural interventions in high-income countries increased in median relative, 22% of cycling behaviour compared to the baseline, Mölenberg et al., 2019).

*Soft* levers are strategies that target a change in psychological factors associated with mobility, such as individual’s intention to use active modes of transport, self-efficacy and attitudes towards active mobility, mobility habits, etc. In the behaviour change literature, these levers refer to behaviour change techniques (Michie, Ashford, et al., 2011). In the field of mobility, some examples of behaviour change interventions are those proposing personalised transport advice, prompting mobility change goal setting and action planning, furnishing maps, transport schedules and other written materials (for reviews, see Arnott et al., 2014; Möser & Bamberg, 2008; Semenescu et al., 2020). The results concerning *soft* levers are more heterogeneous. While a meta-analysis and some systematic reviews show a lack of rigorous studies to allow a conclusion about the effects of these interventions (e.g., Arnott et al., 2014; Graham-Rowe et al., 2011; Semenescu et al., 2020).

While past interventions have shown promising results, most of them share the same limitations. First, few studies rely on “robust” methodologies such as randomised controlled trials comparing the effects of an experimental intervention *vs*. a control intervention (e.g., Graham-Rowe et al., 2011 identified only 14 studies among 77 studies considered as being “robust”). Second, few studies were anchored in behaviour change theories, and few describe the behaviour change techniques that were used to elaborate the intervention (e.g., Aittasalo et al., 2012; Fujii & Taniguchi, 2005; Mutrie et al., 2002). Third, most of the studies use only self-reported measures and no *in situ* sensors to measure behaviour (except from some exceptions like Aittasalo et al., 2019; Ben-Elia & Ettema, 2009). Fourth, few studies have mid- and long-term follow-ups (three months after the intervention or more, e.g., Aittasalo et al., 2012; Cellina et al., 2019; Hemmingsson et al., 2009; Mutrie et al., 2002; Thøgersen, 2009). Fifth, most studies focused on only one type of lever, either *hard* or *soft* (e.g., In their meta-analysis, Semenescu et al., 2020 found only five interventions among 30 studies having combined soft and hard levers). InterMob study aims to address all of these limits.

### 1.2 InterMob Study

InterMob is a study that is part of the interdisciplinary Mobil’Air project that seeks to reduce air pollution in Grenoble, France (Mathy et al., 2020). More precisely, InterMob study combines concepts and methods from geography, psychology, economics, and epidemiology, with the aim to reduce car use and promote active and sustainable mobility in the Grenoble metropolitan area. InterMob seeks to address the aforementioned limitations by being anchored in behaviour change theories (see Figure 1). It also relies on a rigorous methodology (a randomised controlled study comparing an experimental and a control group). In addition, it proposes a longitudinal follow-up (eight weeks of measurement spread over 24 months), the experimental intervention contains *hard* and *soft* levers (e.g., free transport and behaviour change techniques), and it combines *in situ* objective measures and self-reported measurements, in order to collect data on mobility behaviours (e.g., through GPS sensors) as well as on their correlates (e.g., through questionnaires assessing intentions, habits, sociodemographic characteristics, etc.). Moreover, the content of the experimental group is elaborated by mobilising evidence- and theory-based approaches. Indeed, interventions that rely on a theoretical approach appear to be associated with larger effect sizes (Webb et al., 2010) and more consistent effects (McEwan et al., 2019). The active control arm includes sensibilisation to air pollution risks (*soft* lever). It targets attitudes, which have been identified as a factor of behavioral intention (e.g., theory of planned behavior). Such behavior change technique is expected to be less efficient in changing mobility behaviors than the techniques included in the experimental arm, the latter targeting more directly the intention-behavior gap.

**Figure 1.**
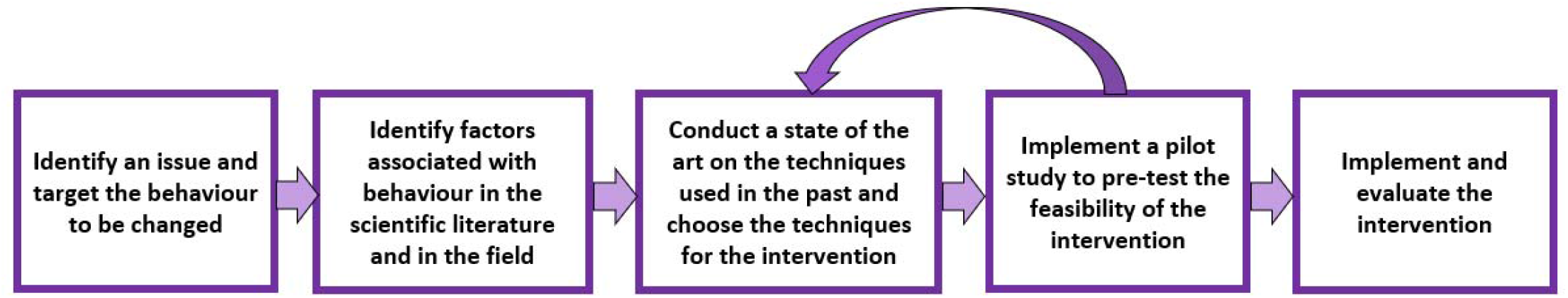
Followed steps for the elaboration, implementation and evaluation of InterMob (the proposed methodology summarises the methodologies proposed by Bartholomew et al., 2016; Bartholomew, 2011; Desrichard et al., 2016; Michie, van Stralen, et al., 2011).

In conclusion, InterMob is a 24-months randomised controlled trial, parallel group, two-arm, superiority trial with 1:1 allocation ratio.

The objectives of the study are to:

a. Evaluate the effectiveness of the InterMob intervention in reducing car use in the short- and in the long-term (one, three, seven, twelve, and twenty-four months after the beginning of the intervention).
b. Identify the mechanisms underlying mobility behaviours and behaviour change (e.g., psychological constructs such as intentions, self-efficacy, habits).
c. Identify the moderators of mobility change (i.e., the conditions and contexts under which the intervention is effective such as family context, degree of cyclability, trip chaining, work situation, activities organisation in space and time).

### 1.3 Procedure for elaborating, implementing, and evaluating a theory- and evidence-base behavioural intervention

To ensure the theory- and evidence-based character of the study, we followed a framework summarising the methodologies and recommendations already used in health psychology (Bartholomew et al., 2016; Desrichard et al., 2016; Hankonen & Hardeman, 2020; Michie, van Stralen, et al., 2011) (see Figure 1).

First, we reviewed the literature on the associations between air pollution, physical inactivity, and mobility behaviours. Then, we undertook a literature review in psychology and geography (e.g., De Witte et al., 2013; Ewing & Cervero, 2010; Lanzini & Khan, 2017; Van Acker et al., 2010) on the factors underlying daily mobility behaviour. Third, we carried out an interdisciplinary literature review to identify the strategies that have been successful in reducing car use and increasing active and sustainable mobility (e.g., Arnott et al., 2014; Bird et al., 2013; Martin et al., 2012; Semenescu et al., 2020). Fourth, we conceived a first version of the InterMob intervention, and conducted a pilot study to test its feasibility. Finally, we established the definitive version of InterMob that is detailed in this article. The present article follows the SPIRIT 2013 Guidance for protocols of critical trials (Chan, Tetzlaff, Altman, et al., 2013; Chan, Tetzlaff, Gotzsche, et al., 2013).

## 2. Methods

### 2.1 Ethics and data protection

The Figure 2 shows the details of the ethics and data protection process. First, InterMob study received the ethic’s approval from Grenoble Alpes Research Ethics Committee (CERGA) in January 2019 (File CER Grenoble Alpes-Avis-2019-01-29-2). Moreover, as part of the application to the Ethics Committee, we carried out a Privacy Impact Assessment (PIA) to assess and avoid any potentially negative impacts of InterMob study on participants’ data privacy. This procedure involved setting a discussion group with the data protection officer of the University, the people in charge of the data infrastructure at the university, and relevant representatives of the target population (members of a car-sharing platform, a local energy and climate agency, and a member of a pedagogical program about air pollution). During the discussion group, we talked about the potential implications of the data-collection tools (e.g., GPS sensors, detailed surveys that include data on psychological constructs as well as the geographic and socio-demographic context), and we detailed the proposed procedures and elements to ensure the respect of the General Data Protection Regulation (GDPR).

**Figure 2.**
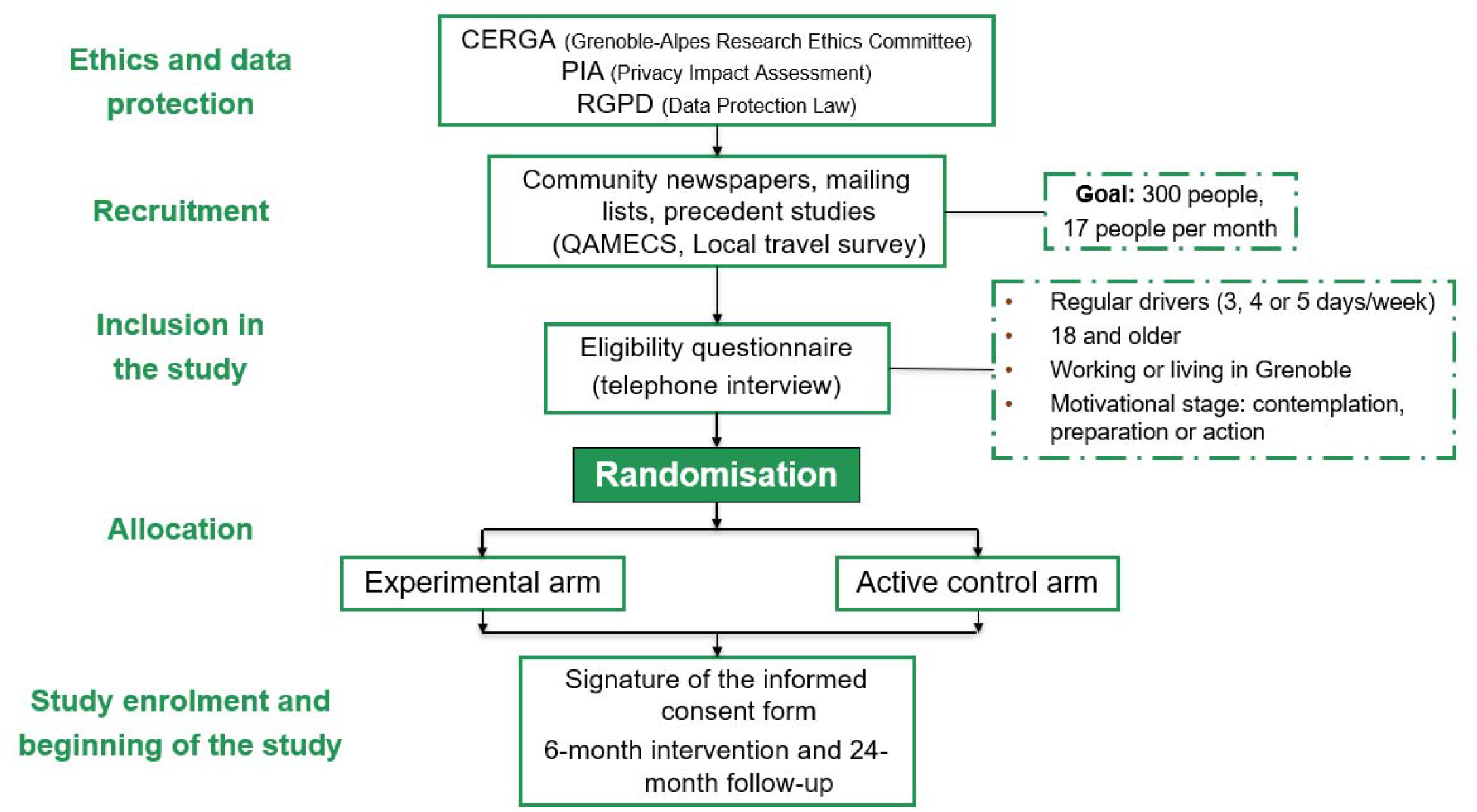
Procedure of the InterMob study since the ethics and data protection procedures to the study enrolment and beginning of the study.

### 2.2 Participants

#### 2.2.1 Study Setting

To detect the effects of InterMob study on car use reduction, recruitment will be restricted to individuals living and working in the Grenoble metropolitan area (see Figure 3), an urban area of south-eastern France (in the Alps) with a population size of approximately 600,000 people. The topography of this area is characterised by being flat in the valleys (especially the Grenoble city) and surrounded by mountains (suburban areas). Car is the main transport mode for daily trips (53% of daily trips in the Grenoble metropolitan area are made by car, SMMAG, 2021).

**Figure 3.**
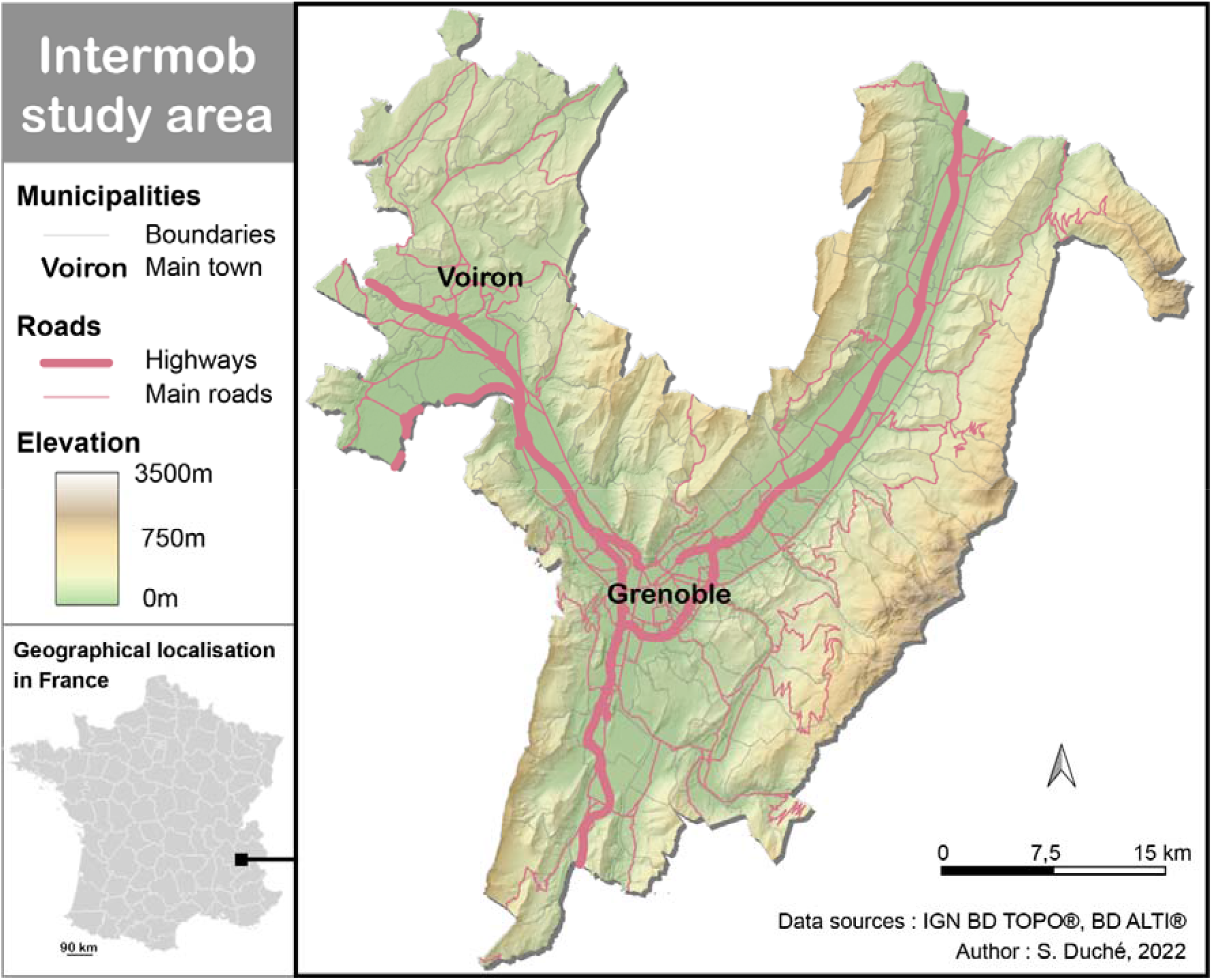
Map of Intemob study area.

#### 2.2.2 Eligibility criteria

To take part in the InterMob study, participants will need to meet the following criteria:

- Being 18 years and older at the time of the eligibility interview;
- Living, working, studying, in the Grenoble metropolitan area;
- Having a car or motorbike as the main transport mode during the week (excluding the weekends);
- Travelling at least three days a week (excluding the weekends) by car or motorbike;
- Motivated to reducing car use for daily trips, or having already started this reduction (i.e., being in the contemplation, preparation or action stages of change according to Prochaska et al., 1994, 2008);
- Expecting to stay in the Grenoble metropolitan area for the duration of the study (24 months).

All the eligibility criteria have been established by considering theoretical and practical aspects. For instance, theories in psychology suggest that participants who intend to change their behaviours do not need the same behavioural techniques than those not having such intention (e.g., Bamberg, 2013; Yang et al., 2010). More precisely, one of the main goals of behaviour change approaches is to help people to overcome the intention-behaviour gap (Sheeran & Webb, 2016).

### 2.3 Participant timeline

#### 2.3.1 Recruitment, allocation, and blinding

The participants will be recruited through different sources: mailing lists from former studies conducted by the research team (QAMECS-SHS and EMC^2^ 2019-2020, local travel surveys, surveys from other laboratories and projects), publicity in local events related to transport or environment, publicity in community newspapers, intervention with the mobility referents of companies in the Grenoble region and, social networks (e.g., Facebook, LinkedIn and Instagram). The people that will be interested to participate in the InterMob study will fill up an online form in order to be contacted by a member of the implementation team (the information in this form includes the first name, a telephone number, an email address, municipality of residence, sex, age, educational attainment and the availability for receiving a phone call). The implementation team will call interested people to explain the study procedure and to administer the eligibility questionnaire. A few days later, the implementation team will call again the eligible people to ask if they accept to participate in the study. If a participant accepts, she/he will be randomly allocated to the experimental or active control arm. For this allocation, a randomisation list for the whole study was created by blocks of six before the beginning of the enrolment (i.e., it exists three possibilities to be in the experimental arm and three possibilities to be in the active control arm) by a member of the scientific team. The list was implemented in the management tool software developed by the team with WebDev for the study. The management tool automatically assigns each eligible participant to a group (the n-th participant is assigned to the group corresponding to the n-th position of the list).

Concerning blinding, the implementation team (programme implementers) will not be aware of group allocation at baseline (S0), but blinding will be impossible afterward, as the programme implementers deliver different contents to each arm through face-to-face interviews with participants. The researchers will not be aware of group allocation at any time, and the researchers responsible for analysing the data will be blinded to the treatment allocation. Double blinding will not be possible given that allocation concealment is impossible for participants in such an intervention.

#### 2.3.2 Timeline of the Study

Participants enrolled in the study will have a first two hours with a programme implementer (“Session 0” or S0, see Figure 4). At the beginning of this meeting, the participant reads and signs an informed consent form (cf. Supplementary materials). She/he fills up a first questionnaire assessing her/his quality of life, biometrics, mobility behaviours, the weekly duration of moderate-to-vigorous physical activity, socio-demographic information and psychological constructs 1, 2 and 3, as well as a mobility logbook assessing her/his last week mobility (for more details about the tools and surveys, see Table 1). Then, the implementation team explains how to correctly carry the Sensedoc™ and MicroPem™ sensors (see Figure 5), and the participant is instructed to start carrying them from the next seven days. At the end of the session, the sensors are collected by the programme implementer. Moreover, during this session, the participants receive a daily survey on their telephones measuring psychological constructs 4. Two weeks after S0, the participants have the first intervention appointment (S0+) that will be described in the intervention subsection.

**Figure 4.**
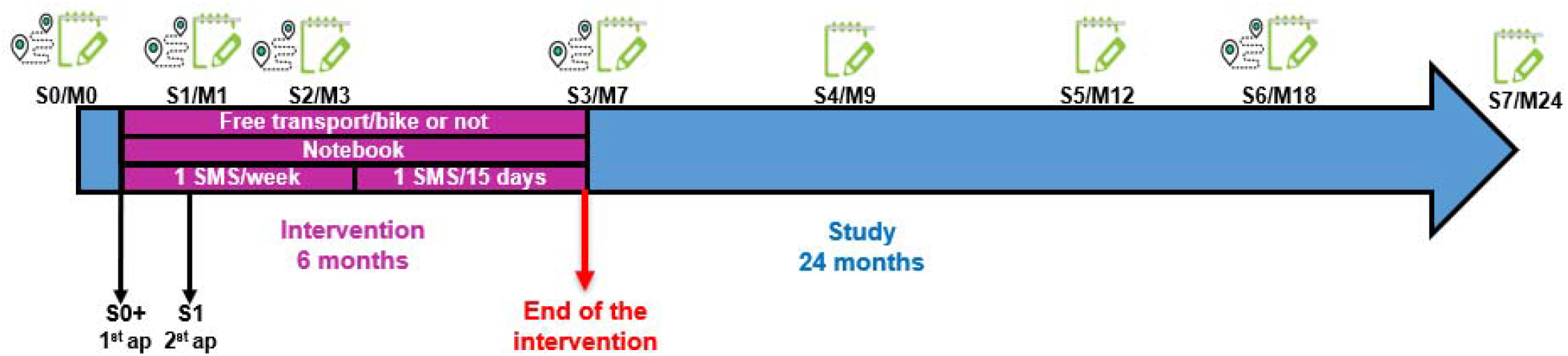
Calendar of the intervention and the measurements. S = Session (seven-day measurement), M = Month, ap= appointment being part of the intervention. 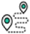 represents seven days of carrying a Sensedoc and a MicroPem (the air pollution sensor is only carried during the sessions 0, 3 and 6), 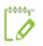 represents seven days of filling up a mobility logbook, answering a long survey (one time) and a short daily survey.

**Table 1.**
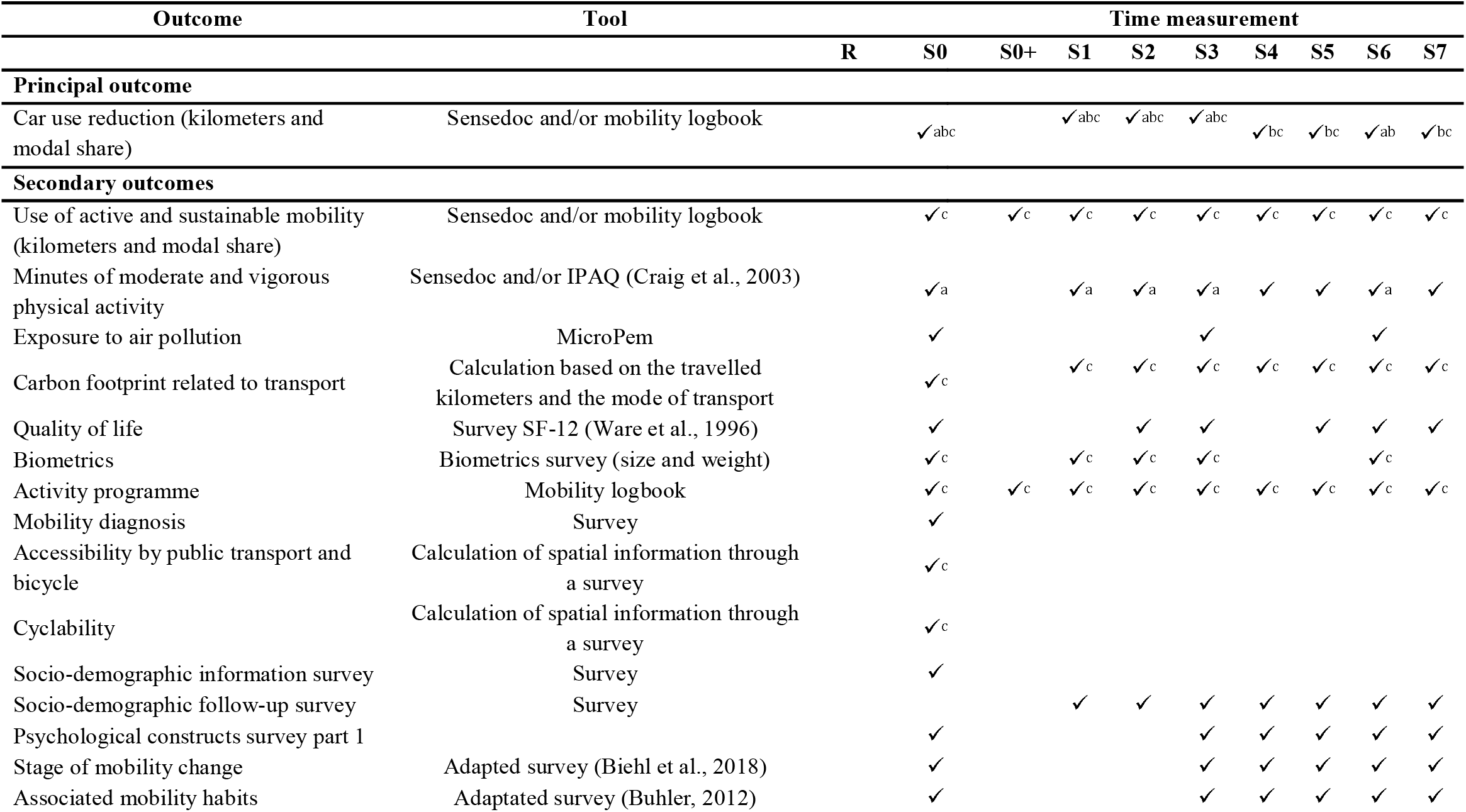
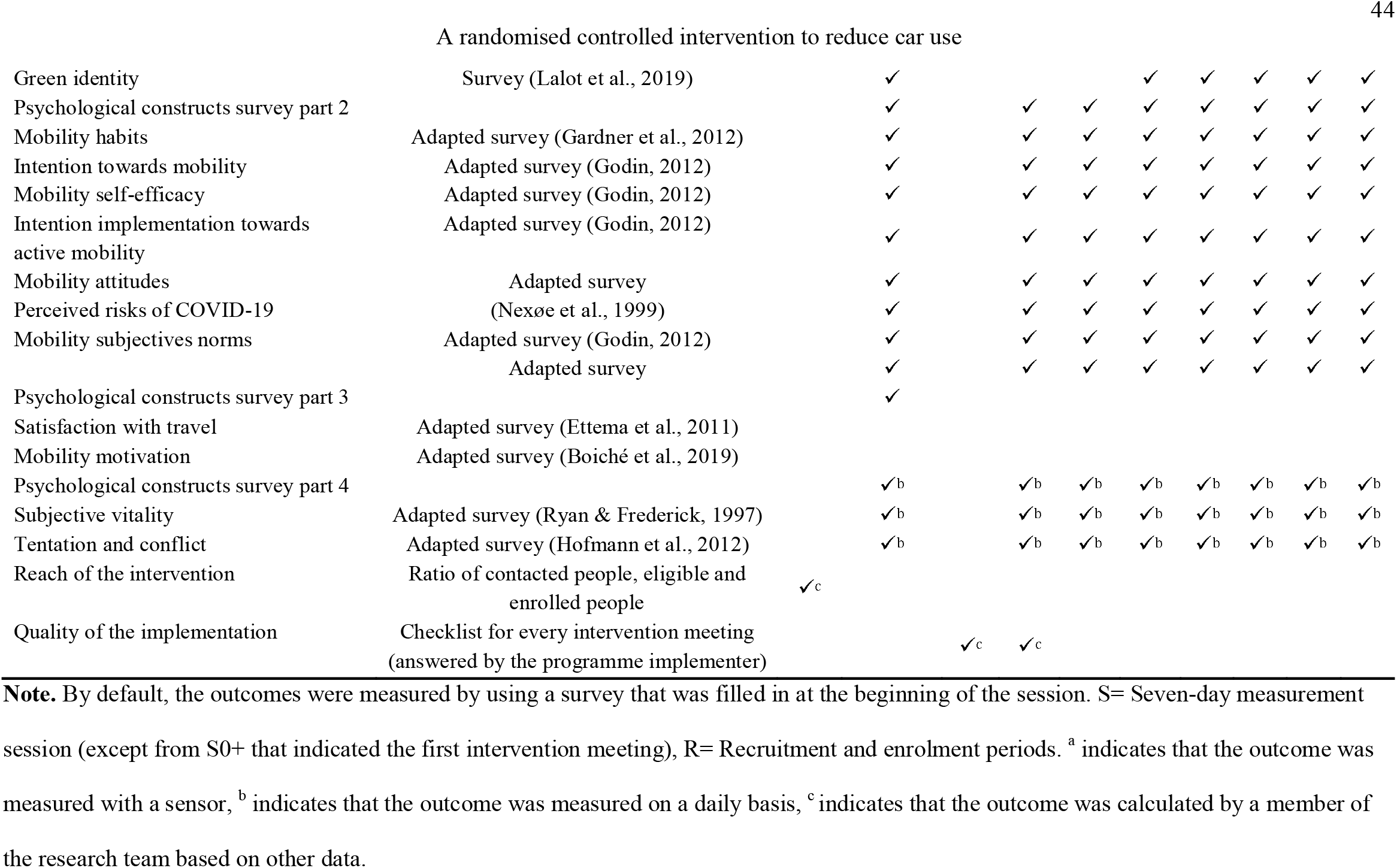
Summary of the outcomes, tools and time measurements.

**Figure 5.**
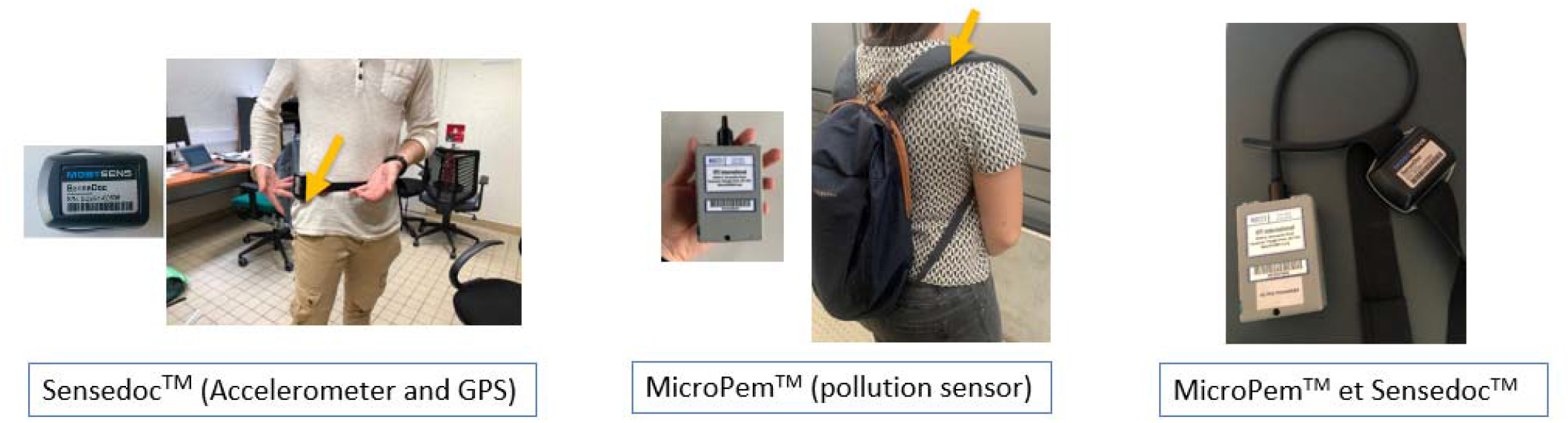
Data collection sensors. In the left side, Sensedoc™ sensor (Accelerometer and GPS) which should be worn in a belt around the waist. In the middle, MicroPem™ (pollution sensor) which should be worn in a bag or purse with the pipe as close to the airway as possible. In the right side, the MicroPem™ and the Sensedoc™.

Approximately one month after the beginning of the study, participants have the second meeting of the intervention (1 hour) and start a new session of measurement (S1). During this session, they carry a Sensedoc™ and answer the same surveys as in S0. Approximately 3.5 months after the start of the study, the participants have the third session (S2) containing the same elements as in S0 except for the pollution sensor. Then, about seven months after the start of the study, the intervention is finished and immediately after the end of the intervention, the participants have the fourth session (S3) during which they carry the Sensedoc™ and the MicroPem™ and answer the same surveys as in S0. Around nine months after the start of the study, participants start the fifth session (S4), they do not carry any sensors but fill up a mobility logbook and answer the same as in S0. Twelve months after the start of the study, participants start the sixth session (S5) with the same protocol as S4. Eighteen months after starting the study, participants start the seventh session (S6) following the same protocol as S0. Finally, 24 months after starting the study, participants start the eighth session (S7) and follow the same protocol as S4 and S5 (see Figure 4).

All the sessions are scheduled with our management tool by considering the availability of the programme implementers. This internal tool is central to implement and chart all steps of the intervention.

#### 2.3.3 Retention of participants

To limit the attrition risk, the implementation team will send a newsletter including information about the study (e.g., recruitment ratios) and the research team (interviews, recommended scientific articles) a few times a year. Moreover, participants taking part in the study until the session 2 will be drawn to win a connected watch.

#### 2.3.4 Power Analysis and Sample Size

Sample size is difficult to estimate when using multilevel modelling as in the present study, given that sample size calculation is sensitive to the values of all the fixed and random parameters included in the models. As such, we estimated the sample size based on the expected effect size of the intervention at the end of the intervention only, which was an expected difference of 17% of the trips made by car at the end of the intervention (Brockman & Fox, 2011) between the experimental and the control group. With a significance level of 0.05 and a statistical power of 80%, the *a priori* power analysis conducted with G*Power 3.1.9.4 (Erdfelder et al., 2009; Faul et al., 2007) indicated that 150 participants per arm (a total of 300 participants) are necessary for finding a difference of 17% between the experimental and the control arms at the end of the intervention. We will therefore recruit participants until exhaustion of our financial and human resources, with this ideal target of 150 participants per arm. Once recruitment is finished, we will compute the minimal statistically detectable effect that can be observed based on the final number of participants (Lakens, 2022).

### 2.4 Interventions

Eligible participants will be randomised in equal proportion to the experimental arm or to the active control group. The duration of both interventions will be the six first months of the 24-month study period (Figure 4). The detailed content of each arm will be described in the next subsections.

#### 2.4.1 Experimental Arm

The participants attributed to the experimental arm will receive a free six-month transport pass and/or a six-month free access to a classic or electric bike (behavioural technique classified as “12.5 Adding objects” according to Michie, Ashford, et al., 2011). They will have two meetings with a programme implementer who will deliver behavioural change techniques, including a discussion about the motivation to change (behavioural techniques known as “motivational interviewing” and “5.2 salience of consequences” according to Hardcastle et al., 2012; Michie, Ashford, et al., 2011), personalised transport advice (behavioural technique classified as “4.1 Instructions on how to perform the behaviour” according to Michie, Ashford, et al., 2011), a mobility change goal setting and action planning (behavioural techniques classified as “1.1 Goal setting” and “1.4 Action planning” according to Michie, Ashford, et al., 2011), and obstacles’ prevention (behavioural technique classified as “1.2 Problem solving” according to Michie, Ashford, et al., 2011).

More precisely, during the first intervention appointment with the programme implementer called “S0+” (1.5 hours), the participant and the programme implementer will discuss about the motivations to change, they will work together on a personalised transport advice considering the participant’s needs, constraints and preferences, the participant will set some change goals and elaborate an action plan, and he/she will examine the possible obstacles to change and how to prevent them. During the second meeting (1 hour, see Figure 4), the programme implementer and the participant will assess and adapt the previously established goals, they will work on an updated personalised transport advice (if the participants ask for it), and on resolving any obstacles they met since the first meeting that have not been resolved.

Following the first meeting of the intervention, participants will be prompt to fill up a “goal notebook” during the six months of the intervention, by setting or reviewing their goals every two weeks, and by taking notes of the experienced obstacles. In addition, they will receive weekly SMS during the first three months of the intervention, and bimonthly SMS during the last three months of the intervention, to a) prompt goal setting and try to keep the same contexts for the goals like the same trips or the same transport modes (behavioural techniques classified as “1.7 Review outcome” and “8.2 Habit formation” according to Michie, Ashford, et al., 2011) and b) prompt self-monitoring related to the consequences of mobility change (e.g., a more important well-being; behavioural technique classified as “2.4 Self-monitoring of outcomes” according to Michie, Ashford, et al., 2011).

#### 2.4.2 Active Control Arm

The participants in the active control arm will also have two meetings with a programme implementer, of the same duration as in the experimental arm. During the first 1.5 hour meeting “S0+”, the programme implementer and the participant will discuss about air pollution (i.e., definition of air pollution sources, population most impacted, air pollution levels in Grenoble, health consequences of pollution and pollution peaks), about the association between air pollution and mobility (i.e., participants and programme implementer discuss about the exposure of car drivers and bike drivers to air pollution), and about the advantages of commuting by car (behavioural technique classified as “5.2 Salience of consequences” according to Michie, Ashford, et al., 2011). During the second 1-hour meeting, the participant and the programme implementer will discuss of the air quality of the last weeks and check if there were some pollution peaks during the last weeks (behavioural technique classified as 5.2 Salience of consequences, according to Michie, Ashford, et al., 2011).

Following the first meeting, participants will be prompt to fill up an “observation notebook” during the six months of intervention, by taking notes of air quality and of every announcement of pollution peak every two weeks. In addition, they will receive weekly SMS during the first three months of the intervention, and bimonthly SMS during the last three months of the intervention, to prompt taking notes of a) air quality and b) any pollution peak announced in the television/radio/telephone (see Table 2 for a summary of the interventions for the experimental and active control arms).

**Table 2.** Summary of the elements of the experimental and the active control arms.

| **Element** | **Experimental arm** | **Active control arm** |
| --- | --- | --- |
| **Material incentive** | 6 months of free transport and/or<br>6 months of free access to a classic or an electric bike |  |
| **2 appointments with a programme implementer** | First appointment: Discussion about the motivations to reduce car use, co-construction of a personalised mobility advice, mobility change goals setting, development of an action plan, prevention of obstacles to change mobility behaviour.<br>Second appointment: Evaluation and adaptation of change goals and resolution of obstacles | First appointment: Concept of air pollution, in-depth questions on pollution (sources of air pollution, health consequences, pollution levels in Grenoble metropolitan area), discussion about the association between air pollution and car use and discussion about the benefits and advantages of the frequent use of the car<br>Second appointment: Evaluation and discussion of the air quality and the identified pollution peaks |
| **Notebook to be filled up** | Goals notebook:<br>1. Setting new goals or adapting goals every two weeks<br>2. Taking notes of the experienced obstacles and looking for solutions | Observation notebook:<br>1. Taking note of the air pollution every two weeks<br>2. Taking note of the announced air pollution peaks (if any) |
| **2 kinds of SMS (1 SMS per week during the first 3 months and 1 SMS bimonthly during the last six months)** | 1. Prompting the Setting and the Adaptation of goals, action planning and, habit formation<br>2. Self-monitoring of positive consequences of mobility change (e.g., better physical state, feeling of well-being) | 1. Taking notes of the air pollution indices and comments<br>2. Taking notes of the announced air pollution peaks and comments |

#### 2.4.3 Adherence

To measure the adherence to each intervention arm, the programme implementer will take notes of the duration of each meeting and of every situation that could disrupt the planned content of the intervention (e.g., participants that do not receive any personalised transport advice). Moreover, one item in the deployed surveys will assess whether the participant sets goals (i.e., Intention implementation towards active mobility survey, see Table 1).

### 2.5 Outcomes and data collection

#### 2.5.1 Primary outcome and data collection methods

The main outcome of InterMob study will be the reduction of car use measured by kilometres travelled by car and the modal share of the car. This outcome will be calculated from segmented and enhanced GPS tracks obtained from a sensor containing a triaxial accelerometer and a continuous GPS tracking MAX-M8 Global Navigation Satellite System receiver from u-blox (Sensedoc™ 2.0, see Figure 5), and by a mobility logbook (i.e., a paper notebook collecting detailed information on the activities and trips made each day, such as departure and arrival times, the address of the arrival point; the transportation mode, the number of people making the trip with the participant). More precisely, The SenseDoc ™ 2.0 acquires GPS data each second et accelerometry with a frequency of 60hz.

#### 2.5.2 Secondary outcomes and data collection methods

Secondary outcomes will include the following variables associated with mobility behaviours: kilometres travelled by active and sustainable mobility, and the modal share of active mobility, physical activity measured by the Sensedoc ™ 2.0 sensor and the IPAQ (i.e., International Physical Activity Questionnaire, Craig et al., 2003), exposure to air pollution (i.e., concentration of fine particles PM_2.5_ and oxidative potential) measured by an air pollution sensor (MicroPem™), activity programme measured by the mobility logbook and, quality of life measured by a survey (Ware et al., 1996) (see Figure 5).

#### 2.5.3 Mediators, moderators, and collection methods

We will investigate whether the effects of the intervention on mobility-related outcomes are mediated by several psychological constructs, including notably attitudes and self-efficacy towards the car (Godin, 2012), active mobility habits (Gardner et al., 2012), motivational stages of mobility change (Biehl et al., 2018) (see Table 1 for all the details), which will be collected by online surveys, using the platform Sphinx iQ2 v 7.3.1.0 (this platform is located at the university to ensure that the data remain in Europe as required by the European data protection law). Self-control constructs, including subjective vitality (Ryan & Frederick, 1997), temptation, conflict, and resistance (Hofmann et al., 2012), will be measured on a daily basis during the data collection weeks, through online surveys on the platform Sphinx iQ2 v 7.3.1.0 as well.

We will also investigate if the efficacy of the intervention depends on several socio-demographic and geographic variables, including the number of children, number of cars, trip chaining, mobility biographies (Müggenburg et al., 2015), cyclability and density of home address and destination address. They will be measured through the data collected by the online surveys, on the platform Sphinx iQ2 v 7.3.1.0.

#### 2.5.4 Reach and Quality of the Implementation

The reach of the intervention (i.e., the number of people contacted by the implementation team, the number of eligible participants, and the enrolled participants) will be assessed by collecting the data from the first contact to the enrolment of participants. Finally, the quality of the implementation will be assessed by analysing a document filled up by the programme implementer at every appointment related to the intervention (i.e., a checklist for every task to be done during the S0+ and S1 appointments).

### 2.6 Data management and statistical methods

#### 2.6.1 Data quality, management, storage, access and confidentiality

In order to monitor the quality of the data, one member of the implementation team and few members of the scientific team checks the correct completion of logbooks, online questionnaires, and the presence of missing data collected by the sensors.

Data from the sensors (Sensedoc™ and MicroPem™) are downloaded after each session to an encrypted computer and stored on a secure and back-up online storage. Data from the online surveys are downloaded twice a month on the same computer. Furthermore, data from all devices (contact forms, sensors, mobility logbooks and online surveys) will be structured and saved in two different blocks stored on distinct servers:

Block 1 will contain the contact file with the names, addresses and contact details of participants for setting the appointments and sending/recovering material. This file will be in the management tool, encrypted and kept separate from other data by the implementation team coordinator. The correspondence table between the code of the participants and their name will also remain in the management tool, a software as a service (SaaS) deployed on a specific server of the university infrastructure accessible only by the implementation team.

Block 2 will contain all the collected data from surveys and sensors. They will be stored locally, on the secured data centre of the university, with access restricted only to InterMob faculty members and implementation team, in line with data protection law. The raw data will be stored by the programme implementers. The data from the GPS/accelerometer and pollution sensors will be downloaded at each return from the field. The survey data, coming from the Sphinx platform, will be regularly uploaded to the storage space. Only implementation team and informatician administrators will have read/write access. The scientific team will only have read access to raw data. Moreover, they have folders with data pre-processing and processing with read/write access, not accessible to the implementation team. Finally, another folder will be created for sharing with external partners.

#### 2.6.2 Data monitoring, harms and auditing

No data monitoring committee has been set up for this study because there is no strong suspicion that the interventions can potentially harm the participants. Nevertheless, there is a risk that some travel modes (e.g., cycling) might lead to traffic accidents. For this reason and in order to protect the participants as much as possible, the helmets will be mandatory throughout the study. At the psychological level, some participants may experience guilt if they do not manage to reduce car use. In order to limit this risk, the team responsible for the implementation insists on a non-guilty and valorising communication and coaching during their face-to-face meetings with the participants (i.e., motivational interviewing). Moreover, participants will be encouraged to share with the implementation team any adverse events associated to mobility change. This information will be stored in a specific document by the coordinator of the implementation of the study.

The coordinator of the implementation of the study and the coordinators of the project have one meeting per week to audit the trial conduct.

#### 2.6.3 Ancillary and post-trial care

No ancillary and post-trial care will be provided.

#### 2.6.4 Statistical methods

##### 2.6.4.1 Analysing the effects of the intervention on primary and secondary outcomes by using multilevel modelling

The analysis of the effectiveness of the intervention on the primary outcome (reduction of car use, see section 2.5.1) and secondary outcomes (mobility-related variables described in section 2.5.2) will be examined at the very short term (three months after the start of the intervention: S2), the short term (seven months after the start of the intervention: S3), the medium term (one year after the start of the intervention: S5) and the long term (18 months after the start of the intervention: S6). More precisely, we will test the differences in car use reduction between the experimental arm and the active control arm, by using multilevel modelling, which is appropriate when data are nested, as it is the case here with several measurement times per participant (e.g., Boisgontier & Cheval, 2016). Moreover, multilevel modelling offers the possibility to analyse incomplete data sets, which is particularly relevant given the duration of the study, which makes likely the occurrence of missing data (Judd et al., 2012). For this purpose, we will follow the steps recommended by Field et al. (2012):

1. Prepare the data by differentiating between- and within-individual constructs.
2. Centre the within-individual variables by participant (i.e., “group centring”).
3. Create a first “constraint” model (a model containing all the random and fixed variables of the final model) and a “null” model including only the “intercept” equal to 1 and only one random variable associated with the identity of the participant (i.e., 1 | Id) to check the intraclass correlation coefficients (ICC).
4. Build the main model.
5. Evaluate the normal distribution of residuals, Cooks distance and influential cases.

The main model will include the following fixed effects: arm, time, and arm x time interaction, with random intercepts for participants and random linear slopes for repeated measures at the participant level.

Moreover, the statistical analysis will follow intention-to-treat principles. This means that once a participant has been randomly assigned to an arm (i.e., experimental or control), his/her data will be included in the analysis (McCoy, 2017). More precisely, even if a participant has not adhered to the intervention or has not finished the study, his/her data will be included to all the analysis.

In addition, we will carry attrition analysis by comparing the characteristics of the participants that drop out the study and the participants that completed the study (e.g., compliance with eligibility criteria, sociodemographic and geographical characteristics).

##### 2.6.4.2 Analysis of the mediatiors of the effects of the intervention

To analyse the mechanisms explaining potential effects of the intervention on car use reduction and mobility-related variables, the investigators will use multilevel mediation models to test the mediating role of psychological constructs (e.g., intention toward active and sustainable mobility, active mobility habits, self-efficacy towards active mobility) in the intervention mobility-related variables relationships.

##### 2.6.4.3 Analysis of the moderators of the effects of the intervention

To analyse the possible moderators of the effects of the intervention on car use reduction and mobility-related variables, the investigators will assess in multilevel models the statistical interactions between socio-demographic, geographic and psychological variables (e.g., number of children, travelled distances, accessibility of home and work, self-control variables) and the allocation to the experimental or control arm. This will allow identifying to what extent geographical, socio-demographic, and psychological variables moderate the effects of the intervention on car use reduction and mobility-related variables.

## 3. Discussion

InterMob study is a 24-month randomised controlled trial aiming to reduce car use of car and motorbike regular drivers. For this purpose, InterMob will include a two-arm intervention of six months: an experimental arm that combines the use of behaviour change techniques previously identified as being successful at changing mobility behaviour (e.g., personalised transport advice, goal setting, action planning) and free access to public transport or/and to a bike; and an active control arm which will target raising awareness about the consequences of air pollution and the link between air pollution and car use.

InterMob will aim to address the limitations of prior behaviour change interventions in the field, including lack of theory-based intervention elaboration, long-term follow-ups, high-quality methodologies (e.g., randomised controlled trial), and *in situ* measures (Arnott et al., 2014; Dixon & Johnston, 2021; Hoffmann et al., 2014; Petrunoff et al., 2016; Yang et al., 2010). By addressing these limitations, InterMob will allow deepening our knowledge of the long-term effects of a theory-based intervention tested in a 2-arm randomised controlled trial that combines *hard* and *soft* levers, with use of *in situ* measures of mobility and correlates.

Furthermore, we will aim to understand the psychological mechanisms that explain the potential effects of the proposed intervention on mobility, as well as the moderating role of geographical, socio-demographic, and psychological factors in this relationship, to better understand the conditions under which the intervention is effective.

Finally, InterMob study will measure the effects of the intervention not only on mobility change, but also on health-related outcomes such as physical activity and exposure to air pollution. Therefore, the results of our study might imply supplementary arguments for governments and politicians to promote car reduction and active and sustainable mobility, and deepen what has been modelled in past studies (Bouscasse et al., 2022).

## Supporting information

SPIRIT guideliness

## Data Availability

The anonymized and aggregated dataset and the codes used to analyse data will be available after the end of the data analysis in the open science framework:

https://osf.io/9anpg/?view_only=240b64a24554468dbc4b5025aca18824

## 4. List of Abbreviations

GPS: Global Positioning System

## 5. Declarations

### 5.1 Ethics approval and consent to participate

InterMob study was approved by Grenoble-Alpes Ethics Committee (File CER Grenoble Alpes-Avis-2019-01-29-2) in January 2019. Moreover, each participant reads and signs a detailed informed consent form (cf. Supplementary materials) if he/she agrees to participate in InterMob study before starting the study.

### 5.2 Consent for publication

All listed authors consented the publication of the manuscript.

### 5.3 Availability of data and material

The anonymized and aggregated dataset and the codes used to analyse data will be available after the end of the data analysis in the open science framework: https://osf.io/9anpg/?view_only=3cba7f109a984abbbc4408fcfa0f8618 with the DOI 10.17605/OSF.IO/9ANPG.

### 5.6 Authors’ contributions

CT-E drafted the manuscript under the supervision of AC and SD. All the authors critically appraised the manuscript, worked on its content, and approved the final version for submission. AC and SC contributed equally to the conception and coordination of this article.

#### 5.6.1 Composition, roles, and responsibilities in the project

AC and SC are the scientific coordinators of the project. LL is the coordinator of the implementation of the study. CT-E elaborated and formulated the details of the intervention under the supervision of AC, SD, and SC. All authors developed the study concept, contributed to improving the study design and data collection, and approved the final version of this study.

##### 5.7 Acknowledgements

The authors thank the IDEX Univ. Grenoble Alpes, the IResP, the Auvergne-Rhône-Alpes Region, the SMAAG and, the Grenoble-Alpes Metropole.

## 6. Trial registration

ClinicalTrials.gov: NCT05096000 on 27/10/2021 (retrospectively registered). https://clinicaltrials.gov/ct2/show/NCT05096000

## 7. Protocol version and protocol amendments

This version of the protocol (2.0) is dated 1 June 2022. Any changes in the protocol will be communicated and shared on ClinicalTrials.gov https://clinicaltrials.gov/ct2/show/NCT05096000

## 8. Dissemination policy

The results of the study will be published in scientific journals and open science platforms. In addition, the results of the study will be written up in a user-friendly style to be sent to all participants and partners of the study after the end of the study.

