## Supplementary material for "InterMob: A 24-month randomised controlled trial comparing the effectiveness of an intervention including behavioural change techniques and free transport versus an intervention including air pollution awareness-raising on car use reduction among regular car users living in Grenoble, France": SPIRIT guideliness

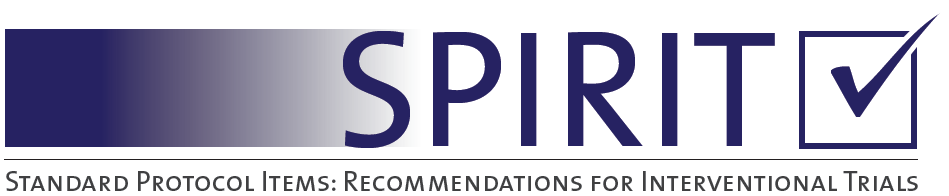

SPIRIT 2013 Checklist: Recommended items to address in a clinical trial protocol and related documents*

**InterMob: A 24-month randomised controlled trial comparing the effectiveness of an intervention including behavioural change techniques and free transport versus an intervention including air pollution awareness-raising on car use reduction among regular car users living in Grenoble, France**

| Section/item | ItemNo | Description |
| --- | --- | --- |
| **Administrative information** | | |
| 🗹Title  Line 1 to 4, page 1 | 1 | Descriptive title identifying the study design, population, interventions, and, if applicable, trial acronym |
| 🗹Trial registration  Line 58, page 4 and lines 569-571, page 25 | 2a | Trial identifier and registry name. If not yet registered, name of intended registry |
|  | 2b | All items from the World Health Organization Trial Registration Data Set |
| 🗹 Protocol version  Lines 572-576 page 25 | 3 | Date and version identifier |
| 🗹 Funding  Lines 547-554 pages 24 | 4 | Sources and types of financial, material, and other support |
| 🗹 Roles and responsibilities  Lines 556-565, page 25 | 5a | Names, affiliations, and roles of protocol contributors |
|  | 5b | Name and contact information for the trial sponsor |
| Line 554-555, page 25 | 5c | Role of study sponsor and funders, if any, in study design; collection, management, analysis, and interpretation of data; writing of the report; and the decision to submit the report for publication, including whether they will have ultimate authority over any of these activities |
| Lines 560-565, page 25 | 5d | Composition, roles, and responsibilities of the coordinating centre, steering committee, endpoint adjudication committee, data management team, and other individuals or groups overseeing the trial, if applicable (see Item 21a for data monitoring committee) |
| 🗹 Background and rationale  Lines 67- 147  Pages 5-8 | 6a | Description of research question and justification for undertaking the trial, including summary of relevant studies (published and unpublished) examining benefits and harms for each intervention |
| Lines 140-147  Page 8 | 6b | Explanation for choice of comparators |
| 🗹Objectives  Lines 150- 158  Page 8 | 7 | Specific objectives or hypotheses |
| 🗹Trial design  Lines 148-149  Page 8 | 8 | Description of trial design including type of trial (eg, parallel group, crossover, factorial, single group), allocation ratio, and framework (eg, superiority, equivalence, noninferiority, exploratory) |
| Methods: Participants, interventions, and outcomes | | |
| 🗹Study setting  Lines 194 – 201  Page 10 | 9 | Description of study settings (eg, community clinic, academic hospital) and list of countries where data will be collected. Reference to where list of study sites can be obtained |
| 🗹 Eligibility criteria  Lines 202 – 220  Pages 10-11  Survey available in Supplemental materials: Lines 323-364, pages 9-10 | 10 | Inclusion and exclusion criteria for participants. If applicable, eligibility criteria for study centres and individuals who will perform the interventions (eg, surgeons, psychotherapists) |
| 🗹Interventions  Lines 303 – 367  Pages 14-17  Table 2, page 45 | 11a | Interventions for each group with sufficient detail to allow replication, including how and when they will be administered |
|  | 11b | Criteria for discontinuing or modifying allocated interventions for a given trial participant (eg, drug dose change in response to harms, participant request, or improving/worsening disease) |
|  | 11c | Strategies to improve adherence to intervention protocols, and any procedures for monitoring adherence (eg, drug tablet return, laboratory tests) |
|  | 11d | Relevant concomitant care and interventions that are permitted or prohibited during the trial |
| 🗹Outcomes  Lines 368 – 409  Pages 17-19  Table 1, pages 43-44 | 12 | Primary, secondary, and other outcomes, including the specific measurement variable (eg, systolic blood pressure), analysis metric (eg, change from baseline, final value, time to event), method of aggregation (eg, median, proportion), and time point for each outcome. Explanation of the clinical relevance of chosen efficacy and harm outcomes is strongly recommended |
| 🗹Participant timeline  Lines 221-299  Pages 11-14 | 13 | Time schedule of enrolment, interventions (including any run-ins and washouts), assessments, and visits for participants. A schematic diagram is highly recommended (see Figure) |
| 🗹Sample size  Lines 288-302  Page 14 | 14 | Estimated number of participants needed to achieve study objectives and how it was determined, including clinical and statistical assumptions supporting any sample size calculations |
| 🗹Recruitment  Lines 222-236  Page 11-12 | 15 | Strategies for achieving adequate participant enrolment to reach target sample size |
| **Methods: Assignment of interventions (for controlled trials)** | | |
| 🗹Allocation:  Lines 234 -242  Pages 11- 12 |  |  |
| 🗹Sequence generation  Lines 236 -239  Page 12 | 16a | Method of generating the allocation sequence (eg, computer-generated random numbers), and list of any factors for stratification. To reduce predictability of a random sequence, details of any planned restriction (eg, blocking) should be provided in a separate document that is unavailable to those who enrol participants or assign interventions |
| 🗹Allocation concealment mechanism  Lines 236 -240  Page 12 | 16b | Mechanism of implementing the allocation sequence (eg, central telephone; sequentially numbered, opaque, sealed envelopes), describing any steps to conceal the sequence until interventions are assigned |
| 🗹Implementation  Lines 236 -242  Page 12 | 16c | Who will generate the allocation sequence, who will enrol participants, and who will assign participants to interventions |
| 🗹Blinding (masking)  Lines 243 - 249  Page 12 | 17a | Who will be blinded after assignment to interventions (eg, trial participants, care providers, outcome assessors, data analysts), and how |
|  | 17b | If blinded, circumstances under which unblinding is permissible, and procedure for revealing a participant’s allocated intervention during the trial |
| **Methods: Data collection, management, and analysis** | | |
| 🗹Data collection methods  Lines 368 - 409  Pages 17-19  Table 1, pages 43-44 | 18a | Plans for assessment and collection of outcome, baseline, and other trial data, including any related processes to promote data quality (eg, duplicate measurements, training of assessors) and a description of study instruments (eg, questionnaires, laboratory tests) along with their reliability and validity, if known. Reference to where data collection forms can be found, if not in the protocol |
| Lines 283-287  Page 14 | 18b | Plans to promote participant retention and complete follow-up, including list of any outcome data to be collected for participants who discontinue or deviate from intervention protocols |
| 🗹Data management  Lines 410-436  Page | 19 | Plans for data entry, coding, security, and storage, including any related processes to promote data quality (eg, double data entry; range checks for data values). Reference to where details of data management procedures can be found, if not in the protocol |
| 🗹Statistical methods  Lines 453 -501  Pages 20-22 | 20a | Statistical methods for analysing primary and secondary outcomes. Reference to where other details of the statistical analysis plan can be found, if not in the protocol |
|  | 20b | Methods for any additional analyses (eg, subgroup and adjusted analyses) |
| Lines 480 - 487  Page 22 | 20c | Definition of analysis population relating to protocol non-adherence (eg, as randomised analysis), and any statistical methods to handle missing data (eg, multiple imputation) |
| **Methods: Monitoring** | | |
| 🗹 Data monitoring  Lines 437 - 450  Page 20 | 21a | Composition of data monitoring committee (DMC); summary of its role and reporting structure; statement of whether it is independent from the sponsor and competing interests; and reference to where further details about its charter can be found, if not in the protocol. Alternatively, an explanation of why a DMC is not needed |
|  | 21b | Description of any interim analyses and stopping guidelines, including who will have access to these interim results and make the final decision to terminate the trial |
| 🗹 Harms  Lines 439 - 448  Page 20 | 22 | Plans for collecting, assessing, reporting, and managing solicited and spontaneously reported adverse events and other unintended effects of trial interventions or trial conduct |
| 🗹 Auditing  Lines 449-450  Page 20 | 23 | Frequency and procedures for auditing trial conduct, if any, and whether the process will be independent from investigators and the sponsor |
| Ethics and dissemination | | |
| 🗹 Research ethics approval  Lines 179 - 193  Page 9-10 and  Lines 529-533  Page 24 | 24 | Plans for seeking research ethics committee/institutional review board (REC/IRB) approval |
| 🗹 Protocol amendments  Lines 571 - 575  Page 25 | 25 | Plans for communicating important protocol modifications (eg, changes to eligibility criteria, outcomes, analyses) to relevant parties (eg, investigators, REC/IRBs, trial participants, trial registries, journals, regulators) |
| 🗹Consent or assent  Lines 252-253  Page 12 and  Lines 531-533  Page 24 | 26a | Who will obtain informed consent or assent from potential trial participants or authorised surrogates, and how (see Item 32) |
|  | 26b | Additional consent provisions for collection and use of participant data and biological specimens in ancillary studies, if applicable |
| 🗹 Confidentiality  Lines 182 – 192 Pages 9-10 and Lines 415 - 436  Pages 19-20 | 27 | How personal information about potential and enrolled participants will be collected, shared, and maintained in order to protect confidentiality before, during, and after the trial |
| 🗹 Declaration of interests  Lines 541 - 545  Page | 28 | Financial and other competing interests for principal investigators for the overall trial and each study site |
| 🗹 Access to data  Lines 432-436  Page 19-20 and  Lines 536-540  Page 24 | 29 | Statement of who will have access to the final trial dataset, and disclosure of contractual agreements that limit such access for investigators |
| 🗹Ancillary and post-trial care  Lines 451 - 452  Page 20 | 30 | Provisions, if any, for ancillary and post-trial care, and for compensation to those who suffer harm from trial participation |
| 🗹Dissemination policy  Lines 576 - 579  Page 25 | 31a | Plans for investigators and sponsor to communicate trial results to participants, healthcare professionals, the public, and other relevant groups (eg, via publication, reporting in results databases, or other data sharing arrangements), including any publication restrictions |
|  | 31b | Authorship eligibility guidelines and any intended use of professional writers |
|  | 31c | Plans, if any, for granting public access to the full protocol, participant-level dataset, and statistical code |
| Appendices |  |  |
| 🗹Informed consent materials  Supplemental materials  Lines 2-322  Page 1 - 8 | 32 | Model consent form and other related documentation given to participants and authorised surrogates |
| Biological specimens  Not applicable | 33 | Plans for collection, laboratory evaluation, and storage of biological specimens for genetic or molecular analysis in the current trial and for future use in ancillary studies, if applicable |

*It is strongly recommended that this checklist be read in conjunction with the SPIRIT 2013 Explanation & Elaboration for important clarification on the items. Amendments to the protocol should be tracked and dated. The SPIRIT checklist is copyrighted by the SPIRIT Group under the Creative Commons “[Attribution-NonCommercial-NoDerivs 3.0 Unported](http://www.creativecommons.org/licenses/by-nc-nd/3.0/)” license.
