## Supplementla Materials for "InterMob: A 24-month randomised controlled trial comparing the effectiveness of an intervention including behavioural change techniques and free transport versus an intervention including air pollution awareness-raising on car use reduction among regular car users living in Grenoble, France"

**Supplemental Material Files**

**Supplemental Material File 1: Infomed consent form – Control group**

**InterMob**
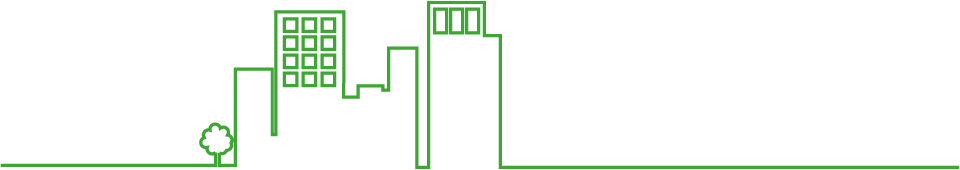

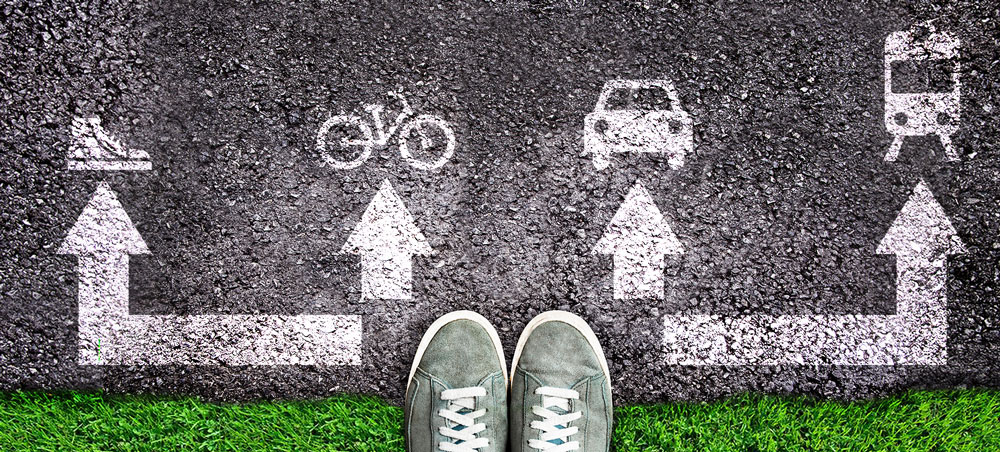

**PA-** **CLEAR CONSENT AND INFORMATION TO THE PARTICIPANT in accordance with the General Data Protection Regulation (GDPR)**

### 1/ The contact details of the Data Protection Officer (DPO) and of the project managers, contacts in case of questions related to the protection of personal data

### Full researchers with scientific responsibility for the project :

| Sonia Chardonnel  Chargée de recherche CNRS  UMR PACTE  14 bis avenue Marie Reynoard,  38100 Grenoble Cedex 9     Téléphone : 04 76 82 20 89 | Aïna Chalabaev  Professeure des Universités  Laboratoire SENS  1741 rue de la piscine  38400 Saint Martin d’Hères     Téléphone : 04 76 63 50 81 |
| --- | --- |

**Data Protection Officer (DPO) :**

**Patrick Guillot**

Data Protection Correspondent (cil)

31, rue des mathématiques - Domaine universitaire
 38400 Saint-Martin-d'Hères

Téléphone : 04 56 52 90 30

### Coordination of the study :

### 2/ The purposes of the processing for which the data are intended

**Aim of the research project: To assess the levers and barriers to change in mobility-related behaviours among people wishing to reduce car use.**

The aim of the InterMob study is to find concrete measures to improve the quality of life of inhabitants of large cities by reducing air pollution and promoting physical activity. This will help in the implementation of measures for public authorities.

### Benefits

The expected benefits of this experiment are to obtain a better understanding of the factors that influence transport mode choices, in order to encourage the use of active modes of travel (e.g. walking, cycling, scootering) and public transport.

**Possible risks**

As far as we know, this research does not involve any risk or discomfort other than that of daily life: you are liable for civil liability (see in your home insurance for example). In case of breakage, loss or theft of the sensors, you will not be held responsible in any way. The university has taken out insurance. InterMob will not be held responsible for any accidents that occur during your travels during this period.

**3/ Third parties who will have access to personal data.**

Your contact details will only be accessible by the survey team in order to plan visits. Other data collected during the study will be anonymised and aggregated with other participants' data for statistical processing by the researchers involved in the study. The processing of your data will take place in France. The data will be stored on a secure European computer system in accordance with Articles 44 to 49 of the GDPR. No data is taken without your knowledge. Thus, we guarantee the respect of your privacy.

**4/ What the processing of your data allows**

This document, "InterMob Informed Consent and Participant Information" signed by you, authorises us to process your data, to store it and to use it for further research on mobility. You have a right of withdrawal, rectification and deletion of your personal data, as explained in points 7 and 8 of this document.

**5/ La durée de conservation des données**

Data will be archived for 15 years from the last date of data collection. As recommended by the Comité d'Éthique en Recherche Grenoble Alpes (CERGA), the consents (necessarily identifiable) will be kept for 10 years from the date of publication and 20 years in case of non-publication, in a sealed envelope in an adapted format bearing the words: "I certify that this envelope contains x (number) consent(s) and x compliant information form(s), collected within the framework of the InterMob study", followed by the name of the scientific manager, Sonia Chardonnel or Aïna Chalabaev.

**6/ What is expected of you**

By agreeing to take part in this research, you will participate in a study on your mobility behaviour, i.e. on the mode(s) of transport you use in your daily life (e.g. car, public transport, bicycle, walking, etc.). The whole study takes place over a total period of 24 months, during 8 sessions spaced out in time (session 0 to session 7), each lasting 7 to 9 days, as well as an appointment called S0+ lasting 1h30. You will receive a call/SMS/email a few days before each session to remind you.

Some sessions require you to wear one or more sensors to collect data related to your activities.

Below is a timeline detailing the monitoring during the study:
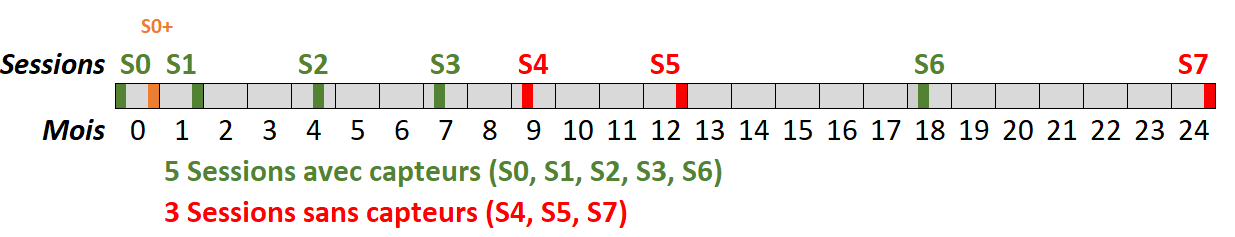

**Interview S0+ :**

The S0+ interview will take place at the beginning of the study, between the S0 and S1 sessions. It will last 1h30 and will consist of a semi-guided interview. The interviewer will give you an observation booklet which you will fill in until the end of our study according to the interviewer's instructions.

**Sequence of sessions :**

**• Sessions with sensor(s) (S0, S1, S2, S3, S6) :**

- On the first day of the session: an interviewer will bring you the sensor(s) and all the documents you will need. He will schedule a link to a short online questionnaire to be sent to you by sms, which you will complete with his assistance. The first session will last 1.5 hours and the other sessions will last about 30 minutes.
- During the 9 days of the session you will have to :
- Wear one or more sensors depending on the session: GPS-accelerometer and pollution sensor (sessions S0, S3, S6), or GPS-accelerometer alone (sessions S1, S2).

The GPS-accelerometer is to be worn on your person, including weekends, from the time you get up to the time you go to bed, except during aquatic activities (showering, bathing, swimming, etc.) or activities that are incompatible with wearing it.

The pollution sensor is to be worn only when you are on the move, and close to you the rest of the time.

These sensors are small boxes that are attached around the waist with an elastic strap for the GPS-accelerometer (the Sensedoc2) or in your bag for the pollution sensor (the MicroPem). They record movements (accelerometer), displacements (GPS) and air pollution. You will be provided with documents to understand the equipment. You can call us at any time.

- Fill in a mobility logbook to indicate each trip made.
- Fill in a short (< 1 min) online questionnaire on your smartphone each evening (link to the questionnaire sent by sms).
- Fill in another online questionnaire received by email on quality of life, physical activities and socio-demographic aspects, your travel habits, your perceptions of the different modes of transport, and your personal situation (duration between 10 and 25 min depending on the session).
- On the last day of the session: an interviewer will collect the sensor(s). You will have to fill in a short online questionnaire about your physical activity during the session (self-administered). If you have worn the pollution sensor during this session (S0, S3, S6), the interviewer will also administer a short questionnaire about the sources of pollution you were exposed to during the session. The appointment will last about 30 minutes.
- **At the first appointment of the first S0 session (RDV S0J0),** you will receive your "welcome pack" with all the necessary documents. You will sign this paper consent after having asked all your questions. You will then be given a presentation of the pollution sensors, the GPS-accelerometer, the mobility logbook, and the tools used to answer questionnaires. With the help of the interviewer, you will fill in a mobility logbook which records your movements of the previous day, and then you will fill in an online questionnaire received by text message. Prior to the meeting, you will have received by email another longer questionnaire to fill in before the meeting, which the interviewer will be able to help you fill in. We will answer your questions so that you can familiarise yourself with the computer support for the questionnaires.

**• Sessions without sensor(s) (S4, S5, S7) :**

- - An interviewer will call you at the beginning and end of the session. The telephone appointments will last about 5 minutes.
- - During the 7 days of the session you will have to :
- Fill in a mobility logbook to indicate each trip made (given at the last face-to-face meeting)
- Fill in a very short online questionnaire each evening on your smartphone received by sms (link to the questionnaire in the sms).
- Fill in an online questionnaire on quality of life, physical activities and socio-demographic aspects, your travel habits, your perceptions of the different modes of transport, and your personal situation (duration between 10 and 25 min depending on the session)
- At the end of the session, you will have to return your mobility booklet by post using a stamped envelope given to you beforehand by one of our interviewers

**SMS :**

You will also receive weekly SMS messages between the S0+ meeting and session 2 (S2) and then fortnightly between sessions 2 (S2) and 3 (S3), for a total of about 6 months. These sms will deliver information related to the topics discussed in your group, and will not require any response from you.

The choice of location for the meetings is up to you: at your home, at your workplace, at the InterMob study premises at Grenoble Alpes University, etc. The dates of the appointments and sessions will be agreed with you from one session to the next.

In view of the COVID19 health situation and depending on the case, the appointments will be made either face-to-face if possible or by videoconference using the Zoom tool. In the latter case, the interviewer will come and deliver to you all the sensors and documents relating to the Study, including this paper consent, before the S0, S0+, S1, S2, S3 and S6 sessions described above. The appointments and/or deliveries will be made in strict compliance with the barrier gestures.

**What matters to us is your participation in our study, regardless of your travel choices.**

**7/ Your rights of access, withdrawal, rectification, deletion, portability (articles 15 and following of the GDPR)^[1]^**

Your contribution to this research is voluntary. Therefore, you may withdraw or stop participating in the survey at any time. Your decision to participate, to refuse to participate, or to stop participating will have no effect on your future relations with the concerned laboratories, and the university. To do so, you just need to contact the InterMob manager by email, specifying in the subject line the right you wish to exercise. For more details, please go to http://bit.ly/InterMob, section "The protection of your data".

**8/ Summary on confidentiality & privacy**

The data obtained will be treated with the utmost confidentiality. You have a randomly generated code for identification. The files including your answers to the questionnaires and the sensor recordings will therefore never include your name and contact details, only the code. No information will be released that could reveal your identity. The data from the sensors and questionnaires will be downloaded to the encrypted computer and then transferred to a secure server at university. Only the scientific managers and assistant researchers will have access to it. You will be able to request the destruction or rectification of your personal data at any time and after the fact, in accordance with the RGPD and the provisions of the French Data Protection Act. You will find more information on the website http://bit.ly/InterMob. You have the contact details of the Data Protection Officer and of the project's scientific manager to assert your rights.

**9/ Consent to participate**

By signing this consent form, you certify that you have read and understood the above information and that we have answered your questions to your satisfaction. You agree to participate in the Intermob study for a period of approximately 24 months, to answer truthfully the questionnaires you will receive and to allow your data to be processed electronically.

Done in duplicate.

Date :

LAST NAME, First name :

Signature preceded by the words « **I willingly agree to participate in the Intermob study** ».

**Supplemental Material File 2: Infomed consent form – Experimental group**

**InterMob**
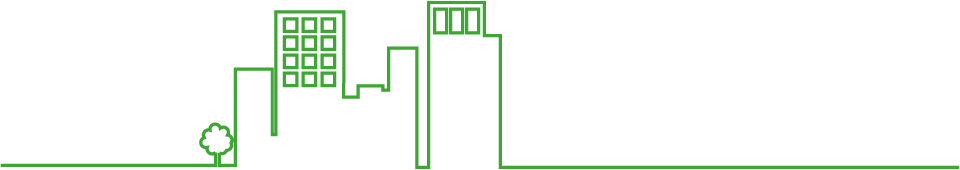

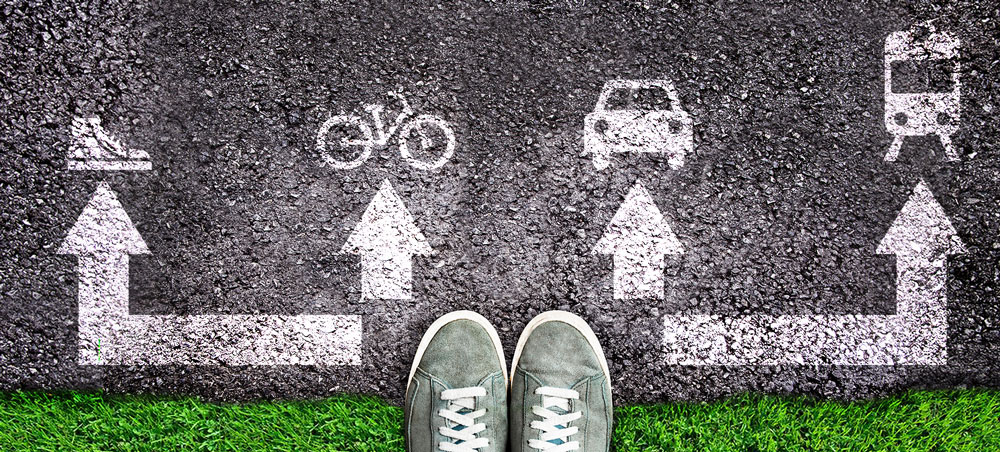

**PB-** **CLEAR CONSENT AND INFORMATION TO THE PARTICIPANT in accordance with the General Data Protection Regulation (GDPR)**

### 1/ The contact details of the Data Protection Officer (DPO) and of the project managers, contacts in case of questions related to the protection of personal data

### Full researchers with scientific responsibility for the project :

| Sonia Chardonnel  Chargée de recherche CNRS  UMR PACTE  14 bis avenue Marie Reynoard,  38100 Grenoble Cedex 9     Téléphone : 04 76 82 20 89 | Aïna Chalabaev  Professeure des Universités  Laboratoire SENS  1741 rue de la piscine  38400 Saint Martin d’Hères     Téléphone : 04 76 63 50 81 |
| --- | --- |

**Data Protection Officer (DPO) :**

**Patrick Guillot**

Data Protection Correspondent (cil)

31, rue des mathématiques - Domaine universitaire
 38400 Saint-Martin-d'Hères

Téléphone : 04 56 52 90 30

Coordination of the study : /

### 2/ The purposes of the processing for which the data are intended

**Aim of the research project: To assess the levers and barriers to change in mobility-related behaviours among people wishing to reduce car use.**

The aim of the InterMob study is to find concrete measures to improve the quality of life of inhabitants of large cities by reducing air pollution and promoting physical activity. This will help in the implementation of measures for public authorities.

### Benefits

The expected benefits of this experiment are to obtain a better understanding of the factors that influence transport mode choices, in order to encourage the use of active modes of travel (e.g. walking, cycling, scootering) and public transport.

**Possible risks**

As far as we know, this research does not involve any risk or discomfort other than that of daily life: you are liable for civil liability (see in your home insurance for example). In case of breakage, loss or theft of the sensors, you will not be held responsible in any way. The University has taken out insurance. InterMob will not be held responsible for any accidents that occur during your travels during this period.

**3/ Third parties who will have access to personal data.**

Your contact details will only be accessible by the survey team in order to plan visits. Other data collected during the study will be anonymised and aggregated with other participants' data for statistical processing by the researchers involved in the study. The processing of your data will take place in France. The data will be stored on a secure European computer system in accordance with Articles 44 to 49 of the GDPR. No data is taken without your knowledge. Thus, we guarantee the respect of your privacy.

**4/ What the processing of your data allows**

This document, "InterMob Informed Consent and Participant Information" signed by you, authorises us to process your data, to store it and to use it for further research on mobility. You have a right of withdrawal, rectification and deletion of your personal data, as explained in points 7 and 8 of this document.

**5/ La durée de conservation des données**

Data will be archived for 15 years from the last date of data collection. As recommended by the Comité d'Éthique en Recherche Grenoble Alpes (CERGA), the consents (necessarily identifiable) will be kept for 10 years from the date of publication and 20 years in case of non-publication, in a sealed envelope in an adapted format bearing the words: "I certify that this envelope contains x (number) consent(s) and x compliant information form(s), collected within the framework of the InterMob study", followed by the name of the scientific manager, Sonia Chardonnel or Aïna Chalabaev.

**6/ What is expected of you**

By agreeing to take part in this research, you will participate in a study on your mobility behaviour, i.e. on the mode(s) of transport you use in your daily life (e.g. car, public transport, bicycle, walking, etc.). The whole study takes place over a total period of 24 months, during 8 sessions spaced out in time (session 0 to session 7), each lasting 7 to 9 days, as well as an appointment called S0+ lasting 1h30. You will receive a call/SMS/email a few days before each session to remind you.

Some sessions require you to wear one or more sensors to collect data related to your activities.

Below is a timeline detailing the monitoring during the study:
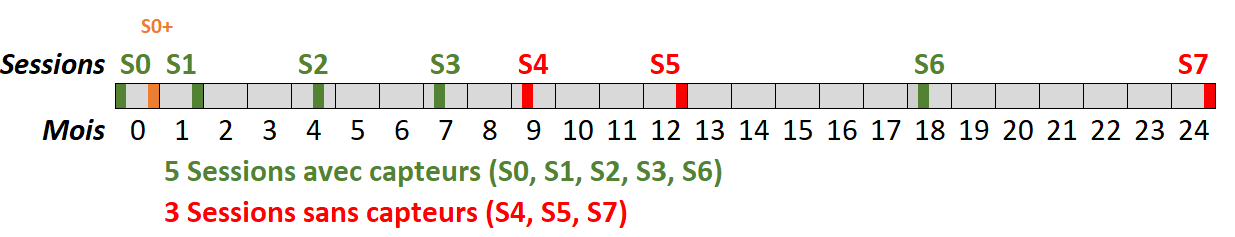

**Interview S0+ :**

The S0+ interview will take place at the beginning of the study, between the S0 and S1 sessions. It will last 1h30 and will consist of a semi-guided interview. The interviewer will give you an objective booklet which you will fill in until the end of our study according to the interviewer's instructions. Eventually, we will need a proof of family income, a copy of your identity card (CNI, passport, driving licence, residence permit), a passport photo, a bank account number, and a copy of your civil disability card (if applicable). Please prepare them now.

**Sequence of sessions :**

**• Sessions with sensor(s) (S0, S1, S2, S3, S6) :**

- On the first day of the session: an interviewer will bring you the sensor(s) and all the documents you will need. He will schedule a link to a short online questionnaire to be sent to you by sms, which you will complete with his assistance. The first session will last 1.5 hours and the other sessions will last about 30 minutes.
- During the 9 days of the session you will have to :
- Wear one or more sensors depending on the session: GPS-accelerometer and pollution sensor (sessions S0, S3, S6), or GPS-accelerometer alone (sessions S1, S2).

The GPS-accelerometer is to be worn on your person, including weekends, from the time you get up to the time you go to bed, except during aquatic activities (showering, bathing, swimming, etc.) or activities that are incompatible with wearing it.

The pollution sensor is to be worn only when you are on the move, and close to you the rest of the time.

These sensors are small boxes that are attached around the waist with an elastic strap for the GPS-accelerometer (the Sensedoc2) or in your bag for the pollution sensor (the MicroPem). They record movements (accelerometer), displacements (GPS) and air pollution. You will be provided with documents to understand the equipment. You can call us at any time.

- Fill in a mobility logbook to indicate each trip made.
- Fill in a short (< 1 min) online questionnaire on your smartphone each evening (link to the questionnaire sent by sms).
- Fill in another online questionnaire received by email on quality of life, physical activities and socio-demographic aspects, your travel habits, your perceptions of the different modes of transport, and your personal situation (duration between 10 and 25 min depending on the session).
- On the last day of the session: an interviewer will collect the sensor(s). You will have to fill in a short online questionnaire about your physical activity during the session (self-administered). If you have worn the pollution sensor during this session (S0, S3, S6), the interviewer will also administer a short questionnaire about the sources of pollution you were exposed to during the session. The appointment will last about 30 minutes.
- **At the first appointment of the first S0 session (RDV S0J0),** you will receive your "welcome pack" with all the necessary documents. You will sign this paper consent after having asked all your questions. You will then be given a presentation of the pollution sensors, the GPS-accelerometer, the mobility logbook, and the tools used to answer questionnaires. With the help of the interviewer, you will fill in a mobility logbook which records your movements of the previous day, and then you will fill in an online questionnaire received by text message. Prior to the meeting, you will have received by email another longer questionnaire to fill in before the meeting, which the interviewer will be able to help you fill in. We will answer your questions so that you can familiarise yourself with the computer support for the questionnaires.

**• Sessions without sensor(s) (S4, S5, S7) :**

- - An interviewer will call you at the beginning and end of the session. The telephone appointments will last about 5 minutes.
- - During the 7 days of the session you will have to :
- Fill in a mobility logbook to indicate each trip made (given at the last face-to-face meeting)
- Fill in a very short online questionnaire each evening on your smartphone received by sms (link to the questionnaire in the sms).
- Fill in an online questionnaire on quality of life, physical activities and socio-demographic aspects, your travel habits, your perceptions of the different modes of transport, and your personal situation (duration between 10 and 25 min depending on the session)
- At the end of the session, you will have to return your mobility booklet by post using a stamped envelope given to you beforehand by one of our interviewers

**SMS :**

You will also receive weekly SMS messages between the S0+ meeting and session 2 (S2) and then fortnightly between sessions 2 (S2) and 3 (S3), for a total of about 6 months. These sms will deliver information related to the topics discussed in your group, and will not require any response from you.

The choice of location for the meetings is up to you: at your home, at your workplace, at the InterMob study premises at the university, etc. The dates of the appointments and sessions will be agreed with you from one session to the next.

In view of the COVID19 health situation and depending on the case, the appointments will be made either face-to-face if possible or by videoconference using the Zoom tool. In the latter case, the interviewer will come and deliver to you all the sensors and documents relating to the Study, including this paper consent, before the S0, S0+, S1, S2, S3 and S6 sessions described above. The appointments and/or deliveries will be made in strict compliance with the barrier gestures.

**What matters to us is your participation in our study, regardless of your travel choices.**

**7/ Your rights of access, withdrawal, rectification, deletion, portability (articles 15 and following of the GDPR)^[2]^**

Your contribution to this research is voluntary. Therefore, you may withdraw or stop participating in the survey at any time. Your decision to participate, to refuse to participate, or to stop participating will have no effect on your future relations with the concerned laboratories, and the University. To do so, you just need to contact the InterMob manager by email, specifying in the subject line the right you wish to exercise. For more details, please go to http://bit.ly/InterMob, section "The protection of your data".

**8/ Summary on confidentiality & privacy**

The data obtained will be treated with the utmost confidentiality. You have a randomly generated code for identification. The files including your answers to the questionnaires and the sensor recordings will therefore never include your name and contact details, only the code. No information will be released that could reveal your identity. The data from the sensors and questionnaires will be downloaded to the encrypted computer and then transferred to a secure server at Grenoble Alpes University. Only the scientific managers and assistant researchers will have access to it. You will be able to request the destruction or rectification of your personal data at any time and after the fact, in accordance with the RGPD and the provisions of the French Data Protection Act. You will find more information on the website http://bit.ly/InterMob. You have the contact details of the Data Protection Officer and of the project's scientific manager to assert your rights.

**9/ Consent to participate**

By signing this consent form, you certify that you have read and understood the above information and that we have answered your questions to your satisfaction. You agree to participate in the Intermob study for a period of approximately 24 months, to answer truthfully the questionnaires you will receive and to allow your data to be processed electronically.

Done in duplicate.

Date :

LAST NAME, First name :

Signature preceded by the words « **I willingly agree to participate in the Intermob study** ».

**Supplemental Material File 3: Eligibility survey**

**QEL_Q01. What is your year of birth?**

**QEL_Q02. Are you currently?**

1. employed/2. Unemployed/3. Retired/4. Student/5. Undergoing work experience or an apprenticeship

**QEL_Q03. In which commune do you live?**

Notes: Grenoble metropolis = 49 communes of the Métro and "extended metropolis" see list.

**QEL_Q03_F01. [If QEL_Q03=other], specify :** ______________________ (text)

**QEL_Q02_F01. [If QEL_Q02=1 or 5] In which commune do you carry out your professional activity?** Drop-down list of communes in the extended metropolis (Métro + extended metropolis #1) + other modalities

**QEL_Q02_F02. [If QEL_Q02_F01=other], specify :** ______________________ (text)

**QEL_Q02_F03. [If QEL_Q02= 4 or 5] In which commune do you attend your studies?**

Drop-down list of communes in the extended metropolis (Métro + extended metropolis #1) + other modalities

**QEL_Q02_F04. [If QEL_Q02_F03=other], specify :** ______________________ (text)

**QEL_Q04. On weekdays (excluding weekends), is the car/motorbike/scooter your main mode of transport as a driver?**

1.Yes/0. No.

**QEL_Q04_F01. [If QEL_Q04=1]How many weekdays (excluding weekends) do you travel by car?**

1 / 2/3/4/5

**QEL_Q04_F02. [If QEL_Q04=1.Yes] You currently use a car/motorcycle/scooter for most of your travel. Which of the following statements best describes you?**

0. I do not intend to reduce the frequency with which I use my car/motorcycle for my usual trips.

1. I am thinking about reducing the frequency with which I use my car/motorcycle for my usual trips.

2. I have already started to reduce the frequency with which I use my car/motorcycle for my usual trips.

**QEL_Q05. Do you expect to live in the Grenoble area in the next two years?**

1.very likely/2. Likely/3. Very unlikely

**QEL_Q05_F01. Do you expect to work in the Grenoble area in the next two years? [If QEL_Q02=1 or 4 or 5]**

1.very likely/2. Likely/3. Very unlikely

**QEL_Q05_F02. [If QEL_Q05 =3 and/or QEL_Q05_F01=3 ] Do you think you live and/or work outside the department of Isère?**

1.Yes/0. No/99. Don't know

**Do you agree to be contacted again for possible participation in this study, as the inclusion criteria of the study extend to your place of residence/workplace?**

1.yes/0. No

**Supplemental Material File 4: Physical activity and full scales of psychological constructs**

**Introduction of every survey**

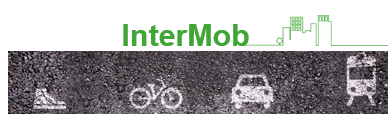

Hello,

We are researchers in geography, urban planning, psychology, economics and epidemiology at the university and we are conducting a research project called InterMob.

This study is carried out in partnership with the university with funding from…

The aim of this questionnaire is to understand the factors that influence mobility behaviour.

We guarantee that the information you give us will be protected in accordance with the General Data Protection Regulation (RGPD). This data will be anonymised before processing and will not be traded in any way. When the data processing is completed, it will be archived at the University for a period of 15 years from the last date of data collection.

For more information on data protection, visit <http://bit.ly/InterMob>

Furthermore, for any type of question concerning the study, the research team can be reached at the following email address:

**QSX_CST. Do you agree to answer the questionnaire?** *

1. yes/0. No

[If QSX_CST = 0. No] Display the following message: End of questionnaire.

[If QSX_CST = 1. Yes] Proceed to the questions corresponding to the session.

**QSX_ID2. Please indicate your participant code.** *

**QSX_ENQ. Is the interviewer present with you?** *

1. yes/0. No

**Physical activity survey**

We are interested in the different types of physical activity you do in your daily life. The following questions ask how much time you spent being physically active in the last 7 days.

Answer each of these questions even if you do not consider yourself to be a physically active person. The questions are about physical activities you do at work, when you are at home, when you travel, and in your free time.

**In the last 7 days, how much time did you spend doing the following behaviours?**

Help: 1h= 60 minutes, 2h = 120 minutes, 3h = 180 minutes, 4h = 240 minutes, 5h=300 minutes, 6h= 360 minutes, 7h = 420 minutes, 8h=480 minutes, 9h= 540 minutes, 10h = 600 minutes

**QAP_Q01. Walking (this includes walking to work and home, walking from one place to another, and any other type of walking you might have done in your free time for relaxation, sport or leisure)**

___ minutes in the last 7 days

**QAP_Q02. Moderate physical activities**

**Moderate physical activities refer to activities that require you to exert moderate physical effort and make it a little harder for you to breathe than normal (for example, carrying light loads, vacuuming, cycling quietly or playing volleyball).**

___ minutes in the last 7 days

**QAP_Q03. Intense physical activities**

**Intense physical activities refer to activities that require you to exert a lot of physical effort and make it much harder for you to breathe than normal (e.g. carrying heavy loads, digging, mountain biking or playing football)**

___ minutes in the last 7 days

**QAP_Q04. Sitting time**

**In the last 7 days, how much time did you spend sitting?**

**This includes time spent sitting at work, at home, when working and in your free time. Examples include sitting at a desk, at a friend's house, reading, sitting or lying down to watch TV, watching a screen.**

__ minutes in the last 7 days

**Psychological constructs survey part 1 : Stage of mobility change, associated mobility habits and green identity**

**Stage of mobility change**

**QL1_Q01. Do you currently use an alternative mode of transport to the car or motorbike (e.g. public transport, walking, cycling) for at least three return trips per week (including weekends)?**

0. I do not use and do not intend to use alternative modes of transport for at least three round trips per week.

1. I am considering using alternative modes of transport for at least three return trips per week

2. I have already started to use alternative modes of transport for at least three round trips per week

**QL1_Q01_F01. [If QL1_Q01. = 0] Do you think that using an alternative mode of transport to the car or motorbike for at least three round trips per week is a realistic option for you?**

1. Yes/0. No

**QL1_Q01_F02. [If QL1_Q01. = 1] Do you plan to use an alternative mode of transport to the car or motorbike for at least three round trips per week in the near future?**

1. Yes/0. No

**QL1_Q01_F03. [If QL1_Q01. = 2] How long ago did you start using an alternative mode of transport to the car or motorbike for at least three round trips per week?**

1. Less than six months/2. Between six and twelve months/3. More than twelve month

**Associated mobility habits**

|  | 3. Always | 2. Often | 1. Rarely | 0. Never |
| --- | --- | --- | --- | --- |
| QL1_Q02. **Listening to music/ radio programme/ ...** |  |  |  |  |
| QL1_Q03. **Thinking about your organisation (work, study, daily life)** |  |  |  |  |
| QL1_Q04. **Reading or rereading documents/Writing a message/SMS** |  |  |  |  |
| QL1_Q05. **Making phone calls** |  |  |  |  |
| QL1_Q06. **Talking/playing with passenger(s)** |  |  |  |  |
| QL1_Q07. **Other (Eating, drinking, smoking, doing hair or make-up)** |  |  |  |  |
| QL1_Q08. **Nothing other than activities necessary for driving** |  |  |  |  |

**Green identity**

**Please qualify the following statements using the following scale:**

| 1  Completely disagree) | 2 | 3 | 4 | 5 | 6 | 7  Completely agree |
| --- | --- | --- | --- | --- | --- | --- |

**QL1_Q09. I consider myself as someone who is interested in environmental issues**

**QL1_Q10. I am a person who supports sustainable development**

**QL1_Q11. I am a person who supports renewable energy**

**QL1_Q12. I see myself as someone who is environmentally conscious**

**QL1_Q13. I consider myself "green".**

**Psychological constructs survey part 2 : Mobility habits, intention towards mobility, Mobility self-efficacy, intention implementation towards active mobility, mobility attitudes, perceived risks of COVID-19, mobility subjectives norms**

**Intention towards mobility**

**Intention to use an alternative means of transport :**

**In the next month for at least three return trips per week (including weekends) ...**

**QL2_Q01. ... do you intend to use a mode of transport other than the car or motorbike?**

1. Not at all/2. Very little intention/3. Somewhat intend/4. Moderately intended/5. Somewhat intention/6. Strongly intend/7. Very strongly the intention

**QL2_Q02. ...will you make an effort to use a mode of transport other than the car or motorbike?** 1.Not at all/2. Very little/3. Somewhat/4. Moderately/5. Somewhat/6. Very much/7. Very strongly

**Habits**

**Car habits**

**Taking the car or motorbike to get around is something that :**

1. Very much disagree/2. Somewhat disagree/3. Slightly disagree/4. Neither disagree nor agree/5. Slightly agree/6. Somewhat agree/7. Strongly agree

**QL2_Q03. ... I do automatically**

**QL2_Q04. ... I do without thinking about it**

**QL2_Q05. ... I can do without paying attention**

**QL2_Q06. ... I start before I even realise it**

**Alternative modes of transport habits**

**QL2_Q07. At present, which alternative modes of transport do you use for your usual trips? (at least once a week)**

(Several answers possible)

1. Walking/2. Cycling (bicycle)/3. EAB - electrically assisted bicycle/5. Public transport (bus, tram, coach)/6. Scooter/7. Electric scooter/8. Other/9. I do not use an alternative mode of transport to the car or motorbike

QL2_Q07_F01. [If QL2_Q07 = 8. other] Specify other main mode of transport: (text)

**[If QL2_Q07 = 1. walking] WALKING IS SOMETHING THAT ... :**

1. Very much disagree/2. Somewhat disagree/3. Slightly disagree/4. Neither disagree nor agree/5. Slightly agree/6. Somewhat agree/7. Strongly agree

**QL2_Q07_F02. ... I do automatically**

**QL2_Q07_F03. ... I do without thinking about it**

**QL2_Q07_F04. ... I can do without paying attention**

**QL2_Q07_F05. ... I start before I even realise it**

**[If QL2_Q07 = 2. Bicycle] Riding a (classic) bicycle IS SOMETHING THAT ... :**

1. Very much disagree/2. Somewhat disagree/3. Slightly disagree/4. Neither disagree nor agree/5. Slightly agree/6. Somewhat agree/7. Strongly agree

**QL2_Q07_F06. ... I do automatically**

**QL2_Q07_F07. ... I do without thinking about it**

**QL2_Q07_F08. ... I can do without paying attention**

**QL2_Q07_F09. ... I start before I have even realised it**

**[If QL2_Q07 = 3. Electrically assisted bicycle] Taking an electrically assisted bicycle IS SOMETHING THAT ... :**

1. Very much disagree/2. Somewhat disagree/3. Slightly disagree/4. Neither disagree nor agree/5. Slightly agree/6. Somewhat agree/7. Strongly agree

**QL2_Q07_F10. ... I do automatically**

**QL2_Q07_F11. ... I do without thinking about it**

**QL2_Q07_F12. ... I can do without paying attention**

**QL2_Q07_F13. ... I start before I have even realised it**

**[If QL2_Q07 = 5. Public transport] TAKING public transport (bus, tram, coach) IS SOMETHING THAT ... :**

1. Very much disagree/2. Somewhat disagree/3. Slightly disagree/4. Neither disagree nor agree/5. Slightly agree/6. Somewhat agree/7. Strongly agree

**QL2_Q07_F14. ... I do automatically**

**QL2_Q07_F15. ... I do without thinking about it**

**QL2_Q07_F16. ... I can do without paying attention**

**QL2_Q07_F17. ... I start before I have even realised it**

**[If QL2_Q07 = 6. scooter] Taking the scooter is something that ... :**

1. Very much disagree/2. Somewhat disagree/3. Slightly disagree/4. Neither disagree nor agree/5. Slightly agree/6. Somewhat agree/7. Strongly agree

**QL2_Q07_F18. ... I do automatically**

**QL2_Q07_F19. ... I do without thinking about it**

**QL2_Q07_F20. ... I can do without paying attention**

**QL2_Q07_F21. ... I start before I even realise it**

**[If QL2_Q07 = 7. electric scooter] Taking the electric scooter IS SOMETHING THAT ... :**

1. Very much disagree/2. Somewhat disagree/3. Slightly disagree/4. Neither disagree nor agree/5. Slightly agree/6. Somewhat agree/7. Strongly agree

**QL2_Q07_F22. ... I do automatically**

**QL2_Q07_F23. ... I do without thinking about it**

**QL2_Q07_F24. ... I can do without paying attention**

**QL2_Q07_F25. ... I start before I even realise it**

**[If QL2_Q07 = 8. other and QL2_Q07_F01 is not empty] Taking 'other 1' IS SOMETHING THAT ...** :

1. Very much disagree/2. Somewhat disagree/3. Slightly disagree/4. Neither disagree nor agree/5. Slightly agree/6. Somewhat agree/7. Strongly agree

**QL2_Q07_F26. ... I do automatically**

**QL2_Q07_F27. ... I do without thinking about it**

**QL2_Q07_F28. ... I can do without paying attention**

**QL2_Q07_F29. ... I start before I even realise it**

**Mobility self-efficacy**

**Self-efficacy towars alternative modes of transport**

**In the coming month, ...**

**QL2_Q08. ... how confident are you that you will be able to use a mode of transport other than the car or motorbike to make at least three round trips per week (including weekends)?**

1.Not at all confident/2. Very little confidence/3. Somewhat confident/4. Moderately confident/5. Somewhat confident/6. Highly confident/7. Very strongly confident

**QL2_Q09. ...how confident are you that you can use an alternative mode of transport to the car or motorbike for at least three return trips per week (including weekends)?**

1.Not at all sure/2. Very unsure/3. Somewhat sure/4. Moderately sure/5. Somewhat certain/6. Very certain/7. Very strongly certain

**QL2_Q10. I think that using an alternative mode of transport to the car or motorbike for at least three return trips per week (including weekends) in the next month is something:**

1.	Very difficult/2. Quite difficult/3. Somewhat difficult/4. Neither difficult nor easy/5. A little easy/6. Quite easy/7. Very easy

**Implementation of intention**

**Over the next month, I have already planned ...**

**QL2_Q11. ... which trips I usually make by car/motorcycle I will replace with another means of transport**

1. Not at all/2. Very little/3. A little/4. Moderately/5. To some extent/6. To a large extent/7. Completely

**QL2_Q12. ... which day of the week/month I will choose to make this trip**

1. Not at all/2. Very little/3. A little/4. Moderately/5. To some extent/6. To a large extent/7. Completely

**QL2_Q13 What alternative mode of transport will I choose to make this trip?**

1. Not at all/2. Very little/3. A little/4. Moderately/5. To some extent/6. To a large extent/7. Completely

**Attitudes**

**QL2_Q14. For me, taking the car or motorbike to make at least three round trips per week (including weekends) in the coming month is...**

1. Very optional/2. Somewhat optional/3. Slightly optional/4. Neither optional nor essential/5. Slightly indispensable/6. Somewhat indispensable/7. Very essential

**QL2_Q15. For me, taking the car or motorbike to make at least three round trips per week (including weekends) during the next month is ...**

1. Very bad/2. Somewhat harmful/3. Slightly harmful/4. Neither harmful nor beneficial/5. Slightly beneficial/6. Somewhat beneficial/7. Very beneficial

**QL2_Q16 For me, taking the car or motorbike to make at least three round trips per week (including weekends) in the next month seems...**

1. Very unpleasant/2. Somewhat unpleasant/3. Slightly unpleasant/4. Neither unpleasant nor pleasant/5. Slightly pleasant/6. Quite pleasant/7. Very pleasant

**QL2_Q17. For me, taking the car or motorbike to make at least three round trips per week (including weekends) during the next month seems...**

1. Very unpleasant/2. Somewhat unpleasant/3. Slightly unpleasant/4. Neither unpleasant nor pleasant/5. Slightly pleasant/6. Somewhat pleasant/7. Very pleasant

**QL2_Q18. For me, taking a mode of transport other than the car or motorbike to make at least three round trips per week (including weekends), over the next month, is...**

1. Very optional/2. Somewhat optional/3. Slightly optional/4. Neither optional nor essential/5. Slightly indispensable/6. Somewhat indispensable/7. Very essential

**QL2_Q19. For me, using a mode of transport other than the car or motorbike to make at least three round trips per week (including weekends) during the next month is...**

1. Very bad/2. Somewhat harmful/3. Slightly harmful/4. Neither harmful nor beneficial/5. Slightly beneficial/6. Somewhat beneficial/7. Very beneficial

**QL2_Q20. For me, taking a mode of transport other than the car or motorbike to make at least three round trips per week (including weekends), over the next month, seems...**

1. Very unpleasant/2. Somewhat unpleasant/3. Slightly unpleasant/4. Neither unpleasant nor pleasant/5. Slightly pleasant/6. Quite pleasant/7. Very pleasant

**QL2_Q21. For me, taking a mode of transport other than the car or motorbike to make at least three round trips per week (including weekends), during the coming month, seems...**

1. Very unpleasant/2. Somewhat unpleasant/3. Slightly unpleasant/4. Neither unpleasant nor pleasant/5. Slightly pleasant/6. Somewhat pleasant/7. Very pleasant

**Mobility subjectives norms**

**Most people who are important to me (family, friends, colleagues) ...**

1.very much disagree/2. Somewhat disagree/3. Slightly disagree/4. Neither disagree nor agree/5. Slightly agree/6. Somewhat agree/7. Strongly agree

**QL2_Q22. ... encourage me to use the car for at least three round trips per week (including weekends)**

**QL2_Q23. ... think that I should use the car to make at least three round trips per week (including weekends)**

**QL2_Q24. ... encourage me to use a mode of transport other than the car to make at least three round trips per week (including weekends)**

**QL2_Q25. ... think that I should use a mode of transport other than the car to make at least three round trips per week (including weekends)**

**QL2_Q26. When I regularly use the car to make at least three round trips per week (including weekends), most people who are important to me (family, friends, colleagues) ...**

Disagree very strongly/2. Disagree somewhat/3. Disagree slightly/4. Neither disapprove nor approve/5. Slightly approve/6. Approve somewhat/7. Strongly approve

**QL2_Q27. When I use a mode of transport other than the car for at least three round trips per week (including weekends), most people who are important to me (family, friends, colleagues) ...**

Disagree very strongly/2. Disagree somewhat/3. Disagree slightly/4. Neither disapprove nor approve/5. Slightly approve/6. Approve somewhat/7. Strongly approve

**The proportion of people in my circle who take ...**

**QL2_Q28. ... the car or motorbike to make at least three return trips per week (including weekends) is**

1. No one/2. A quarter (25%)/3. Half/4. Three quarters (75%)/5. All people

**QL2_Q29. ... transport other than the car to make at least three return trips per week (including weekends) is**

1. No one/2. A quarter (25%)/3. Half/4. Three quarters (75%)/5. All

**Perceived risks of COVID-19**

**Perceived vulnerability**

**The following questions ask about your perceptions of the current health crisis.**

1.Very much disagree/2. Somewhat disagree/3. Slightly disagree/4. Neither disagree nor agree/5. Slightly agree/6. Somewhat agree/7. Strongly agree

**QL2_Q30. I have a high risk of catching the coronavirus disease**

**QL2_Q31. I am concerned about the risk of getting the coronavirus**

**QL2_Q32. I get sick more easily than other people my age**

**Perceived severity**

**QL2_Q33. Getting the coronavirus could cause me serious health problems**

**QL2_Q34. I am afraid that the coronavirus will make me very ill**

**QL2_Q35. I could not bear to get the Coronavirus because of my general health**

**Psychological constructs survey part 3 : Satisfaction with travel, mobility motivation**

**Satisfaction with travel**

**In general, when you are in a car or on a motorbike, how do you feel? Please consider the car (or motorbike for motorcyclists) and give us your opinion on the items listed below:**

|  | **-4** | **-3** | **-2** | **-1** | **0** | **1** | **2** | **3** | **4** |
| --- | --- | --- | --- | --- | --- | --- | --- | --- | --- |
| **QL3_Q01. I am in a hurry -4 to 4 Relaxed** |  |  |  |  |  |  |  |  |  |
| **QL3_Q02. I am afraid of not being on time -4 to 4 sure I will be on time** |  |  |  |  |  |  |  |  |  |
| **QL3_Q03. I am stressed -4 to 4 Calm** |  |  |  |  |  |  |  |  |  |
| **QL3_Q04. I am tired -4 to 4 Awake** |  |  |  |  |  |  |  |  |  |
| **QL3_Q05. I am bored -4 to 4 Enthusiastic** |  |  |  |  |  |  |  |  |  |
| **QL3_Q06. I am bored -4 to 4 Interested** |  |  |  |  |  |  |  |  |  |
| **QL3_Q07. This is the worst -4 to 4 best mode of transport I can think of** |  |  |  |  |  |  |  |  |  |
| **QL3_Q08. This mode of transport is poor quality -4 to 4 very good quality** |  |  |  |  |  |  |  |  |  |
| **QL3_Q09. This mode of transport works very badly -4 to 4 works very well** |  |  |  |  |  |  |  |  |  |
| **QL3_Q10. With this mode of transport I have no risk at all -4 to 4 I have a high risk of catching COVID-19** |  |  |  |  |  |  |  |  |  |

**Please consider public transport and give us your views on the items listed below:**

|  | **-4** | **-3** | **-2** | **-1** | **0** | **1** | **2** | **3** | **4** |
| --- | --- | --- | --- | --- | --- | --- | --- | --- | --- |
| **QL3_Q01. I am in a hurry -4 to 4 Relaxed** |  |  |  |  |  |  |  |  |  |
| **QL3_Q02. I am afraid of not being on time -4 to 4 sure I will be on time** |  |  |  |  |  |  |  |  |  |
| **QL3_Q03. I am stressed -4 to 4 Calm** |  |  |  |  |  |  |  |  |  |
| **QL3_Q04. I am tired -4 to 4 Awake** |  |  |  |  |  |  |  |  |  |
| **QL3_Q05. I am bored -4 to 4 Enthusiastic** |  |  |  |  |  |  |  |  |  |
| **QL3_Q06. I am bored -4 to 4 Interested** |  |  |  |  |  |  |  |  |  |
| **QL3_Q07. This is the worst -4 to 4 best mode of transport I can think of** |  |  |  |  |  |  |  |  |  |
| **QL3_Q08. This mode of transport is poor quality -4 to 4 very good quality** |  |  |  |  |  |  |  |  |  |
| **QL3_Q09. This mode of transport works very badly -4 to 4 works very well** |  |  |  |  |  |  |  |  |  |
| **QL3_Q10. With this mode of transport I have no risk at all -4 to 4 I have a high risk of catching COVID-19** |  |  |  |  |  |  |  |  |  |

**Please consider the bike and give us your opinion on the items listed below:**

|  | **-4** | **-3** | **-2** | **-1** | **0** | **1** | **2** | **3** | **4** |
| --- | --- | --- | --- | --- | --- | --- | --- | --- | --- |
| **QL3_Q01. I am in a hurry -4 to 4 Relaxed** |  |  |  |  |  |  |  |  |  |
| **QL3_Q02. I am afraid of not being on time -4 to 4 sure I will be on time** |  |  |  |  |  |  |  |  |  |
| **QL3_Q03. I am stressed -4 to 4 Calm** |  |  |  |  |  |  |  |  |  |
| **QL3_Q04. I am tired -4 to 4 Awake** |  |  |  |  |  |  |  |  |  |
| **QL3_Q05. I am bored -4 to 4 Enthusiastic** |  |  |  |  |  |  |  |  |  |
| **QL3_Q06. I am bored -4 to 4 Interested** |  |  |  |  |  |  |  |  |  |
| **QL3_Q07. This is the worst -4 to 4 best mode of transport I can think of** |  |  |  |  |  |  |  |  |  |
| **QL3_Q08. This mode of transport is poor quality -4 to 4 very good quality** |  |  |  |  |  |  |  |  |  |
| **QL3_Q09. This mode of transport works very badly -4 to 4 works very well** |  |  |  |  |  |  |  |  |  |
| **QL3_Q10. With this mode of transport I have no risk at all -4 to 4 I have a high risk of catching COVID-19** |  |  |  |  |  |  |  |  |  |

**Mobility motivation**

**Self-determined motivation to take a car:**

**The main reason I intend to use a car or motorbike for my regular travel is...**

1. Very much disagree/2. Somewhat disagree/3. Slightly disagree/4. Neither disagree nor agree/5. Slightly agree/6. Somewhat agree/7. Strongly agree

**QL3_Q31. ...for the pleasure of driving [Intrinsic motivation]**

**QL3_Q32. ...because I would feel ashamed to use other modes of transport [Introjected motivation] QL3_Q33.**

**QL3_Q33. ... because I really believe it is important to take the car [Identified motivation] QL3_Q34. ... because I really believe it is important to take the car [Identified motivation] QL3_Q35.**

**QL3_Q34. ... because people around me urge me to take the car [External motivation].**

**QL3_Q35. ... because other people around me do not like me to use other modes of transport. [External motivation]**

**QL3_Q36. ... because I would feel guilty about not taking the car [Introjected motivation].**

**QL3_Q37. ... because I do it without hesitation [Identified motivation]**

**QL3_Q38. ... because I like it. [Intrinsic motivation]**

**QL3_Q39. ... because it is an integral part of my life [Integrated motivation].**

**QL3_Q40. ... because it is part of the way I have chosen to live my life. [Integrated Motivation]**

**Self-determined motivation to take alternative transport :**

**Table presentation of questions QL3Q60 to QL3Q69**

**If I intend to use an alternative mode of transport to the car or motorbike for my usual trips, it is mainly because...**

1. Very much disagree/2. Somewhat disagree/3. Slightly disagree/4. Neither disagree nor agree/5. Slightly agree/6. Somewhat agree/7. Strongly agree

**QL3_Q41. ... for the pleasure of using other modes of transport as alternatives to the car [Intrinsic motivation] QL3_Q42.**

**QL3_Q42. ... because I would feel ashamed if I didn't do it [Introjected motivation]**

**QL3_Q43. ... because I really believe that it is important to take alternative modes of transport to the car [Identified motivation].**

**QL3_Q44. ... because people around me urge me to use alternative modes of transport to the car [External motivation].**

**QL3_Q45. ... because other people around me do not like the fact that I take the car for most of my trips. [External motivation]**

**QL3_Q46. ... because I would feel guilty about not taking alternative modes of transport**

**QL3_Q47. ... because I would do it without hesitation [Identified motivation]**

**QL3_Q48. ... because I like it. [Intrinsic motivation]**

**QL3_Q49. ... because it is an integral part of my life. [Integral motivation].**

**QL3_Q50. ... because it is part of the way I have chosen to live my life. [Integrated motivation]**

**Psychological constructs survey part 4 : Subjective vitality, tentation and conflict**

**Subjective vitality**

**Tentation and conflict**

**1.	Did you plan to use an alternative mode of transport today? QL4Tentation_behaviour**

1./Yes/2./No

**2.	Did you want to use your car today? QL4Tentation1**

1./Yes/2./No

**3.	How strong was the desire? QL4Tentation_strong**

1./ Very weak desire.............. 7. Irresistible desire

**4.	How much did this temptation conflict with your long-term personal intentions/goals was the desire strong? QL4Tentation_Conflict**

1./ Very low conflict.............. 7. Very high conflict

**5.	How hard did you try to resist/are you resisting this desire? QL4Tentation_resistanceforce**

1 / Not at all resisting.............. 7. Very strongly resist

**6.	Were you able to resist the temptation? QL4Tentation_resistucces**

**1./Yes/2./No**

**Subjective vitality**

**7.	During the day, did you feel full of vitality, in great shape? QL4Vitality1**

1./Not at all..... 7./At all

**8. During the day, did you feel energetic, lively? QL4Vitality2**

1./Not at all..... 7./At all

1. https://www.cnil.fr/fr/reglement-europeen-protection-donnees/chapitre3#Article15
2. https://www.cnil.fr/fr/reglement-europeen-protection-donnees/chapitre3#Article15
